# Automatic cerebral angiographic perfusion analysis

**DOI:** 10.64898/2026.11.24.26368862

**Authors:** Lily Pwint Thinzar, Nanthiya Sujijantarat, Ryan Hebert, Charles Matouk, David Y Chung, Ona Wu, Nils H Petersen, Kyle A Lyman

## Abstract

**Purpose:** To automate anatomical localization for angiographic perfusion and evaluate agreement with manual processing between biplane projections in a held-out external cohort.

**Methods:** Separate anteroposterior (AP) and lateral projection hemisphere and internal carotid artery (ICA) models used DeepLabv3+ with ResNet50 encoders. Development used patient-separated training, validation, and testing. External patients were excluded from fitting and model selection. Automated pipeline and manual-pipeline perfusion shared downstream calculations. Eigible acquisitions contained at least 7 seconds of data after bolus arrival. Segmentation metrics and Bland–Altman analyses evaluated mean transit time (MTT) and time to maximum of the residue function (Tmax).

**Results:** External segmentation testing included 353 runs from 73 patients. Patient-mean hemisphere Dice was 0.959 (95% CI, 0.952-0.965) in AP and 0.965 (0.960-0.969) in lateral views while ICA Dice was 0.635 (0.617-0.650) and 0.558 (0.534-0.579). Manually processed perfusion comparisons included 165 external runs from 52 patients. Mean absolute differences (95% CI) were 0.39 (0.31-0.47) and 0.32 (0.25-0.40) seconds for AP and lateral MTT, and 0.23 (0.15-0.35) and 0.31 (0.17-0.51) seconds for Tmax. Among 220 external cohort biplane pairs from 61 patients, mean absolute differences were 0.48 seconds (95% CI, 0.42–0.53) for MTT and 0.53 seconds (0.35–0.75) for Tmax. Repeated-measures mean biases (95% CI) were −0.07 (−0.19 to 0.05) and 0.16 seconds (−0.08 to 0.40), with 95% limits of agreement of −1.38 to 1.24 and −2.65 to 2.97 seconds respectively.

**Conclusion:** Our automated perfusion pipeline showed favorable agreement with manually processed results in the external validation cohort. Independently processed biplane views had small average bias.

## Introduction

Delayed cerebral ischemia (DCI) remains a major challenge after aneurysmal subarachnoid hemorrhage (aSAH) [1–3]. Clinical assessment, vascular imaging, and cross-sectional perfusion imaging address different aspects of its pathophysiology [4–6]. Digital subtraction angiography (DSA) depicts vascular anatomy during endovascular treatment, but arterial narrowing alone does not describe the downstream tissue response [7, 8]. Quantitative perfusion derived from the angiogram could characterize physiology when treatment decisions are made.

Angiographic perfusion methods estimate tissue transit from contrast passage [8–13]. Our previous work linked these measurements to rescue-therapy responses after aSAH [8], and introduced gamma-variate extrapolation to reduce acquisition-truncation errors [11, 12]. However, the pipeline required manual processing and analyzed lateral images exclusively [8, 11, 12]. We sought to automate hemispheric region-of-interest (ROI) and arterial input function (AIF) localization and extend processing to anteroposterior (AP) images in an automatic workflow.

We developed separate AP and lateral projection hemisphere and internal carotid artery (ICA) segmentation models. Evaluation comprised independent segmentation testing, same-acquisition comparison with manually processed perfusion results, and comparison of independently processed biplane views (Fig. 1).

**Figure 1.**
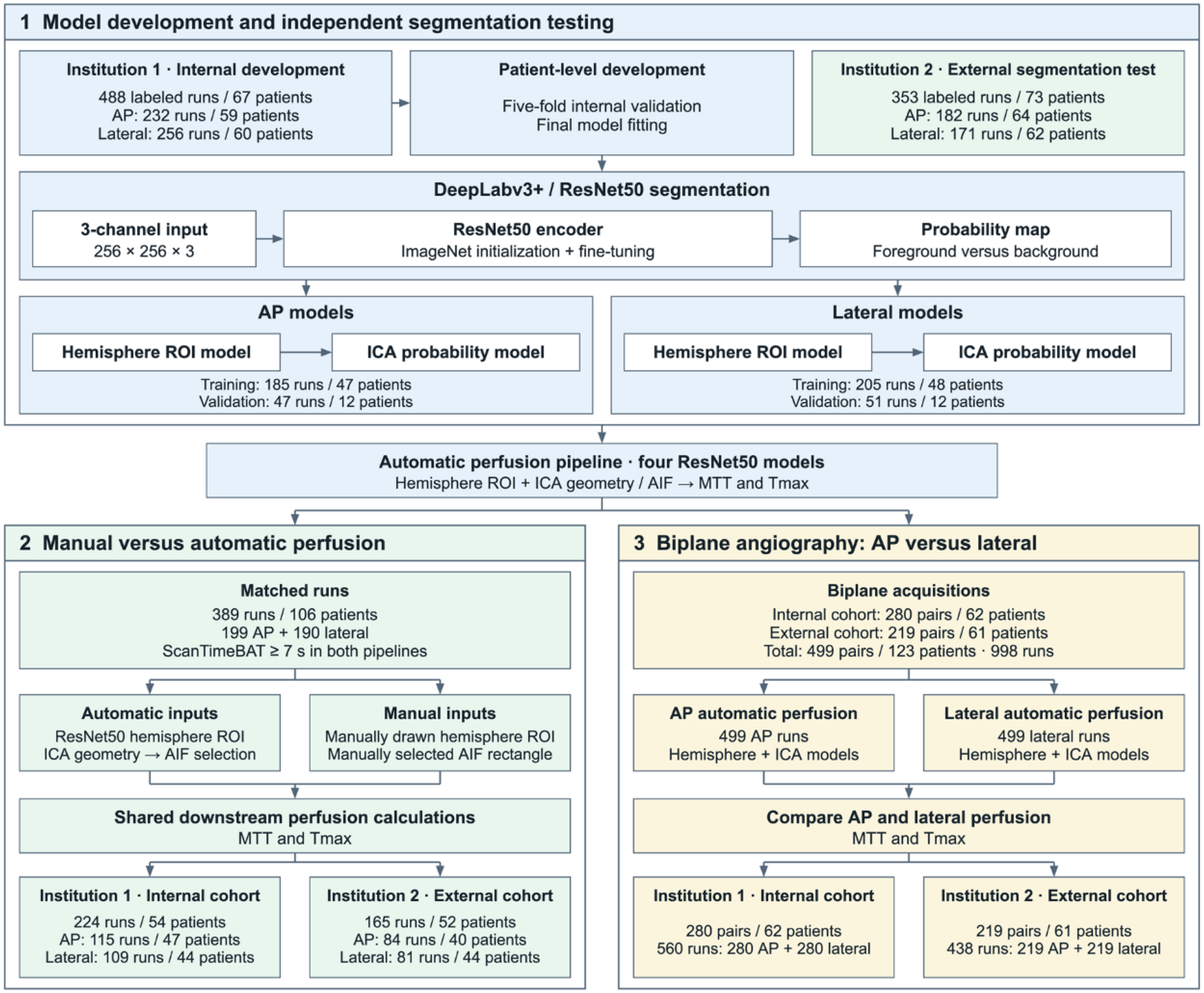
Study design and evaluation cohorts. Three phases link patient-separated model development to manual-input and biplane perfusion comparisons. Segmentation uses 488 internal runs/67 patients and 353 external runs/73 patients. Primary perfusion analyses include 165 external manual-comparison runs/52 patients and 220 external biplane pairs/61 patients. Each biplane pair contains two projection-specific runs.

## Methods

### Study population and evaluation cohorts

This retrospective study included adults with angiographically confirmed aSAH undergoing endovascular rescue therapy. Institutional review boards at Massachusetts General Hospital and Yale New Haven Hospital approved the study with waivers of informed consent, hereafter referred to anonymously as Institution 1 and 2. Eligible admission periods were June 2017–April 2023 at Institution 1 and April 2019–February 2025 at Institution 2. Institution 1 provided the internal validation cohort for development and patient-separated testing while Institution 2 provided the external validation cohort. All DSA data acquired during routine clinical care were considered. An angiography run denotes one projection image series during one injection of contrast while simultaneous AP–lateral biplane image acquisitions yield two runs [14]. Online Resource 1 contains cohort demographics and acquisition characteristics (Tables S1–S2).

### Reference annotations and image channels

One analyst (KL) annotated hemisphere boundaries and ICA centerlines in MATLAB R2025a using randomized acquisition identifiers and site blinding. Reference masks used the uncropped 512 × 512 grid. Lateral contours followed the ophthalmic artery takeoff and superior sagittal sinus to the torcula, with a closing segment [8, 11, 12]. AP hemispheric contours followed cortical venous and sagittal/transverse sinus boundaries (Fig. 2). ICA centerlines extended from the inferior image boundary to the ophthalmic takeoff laterally or skull base in AP views. Training and evaluation targets dilated each ICA centerline by a radius-two-pixel disk. ICA Dice therefore measures narrow-region overlap rather than one-pixel centerline overlap.

**Figure 2.**
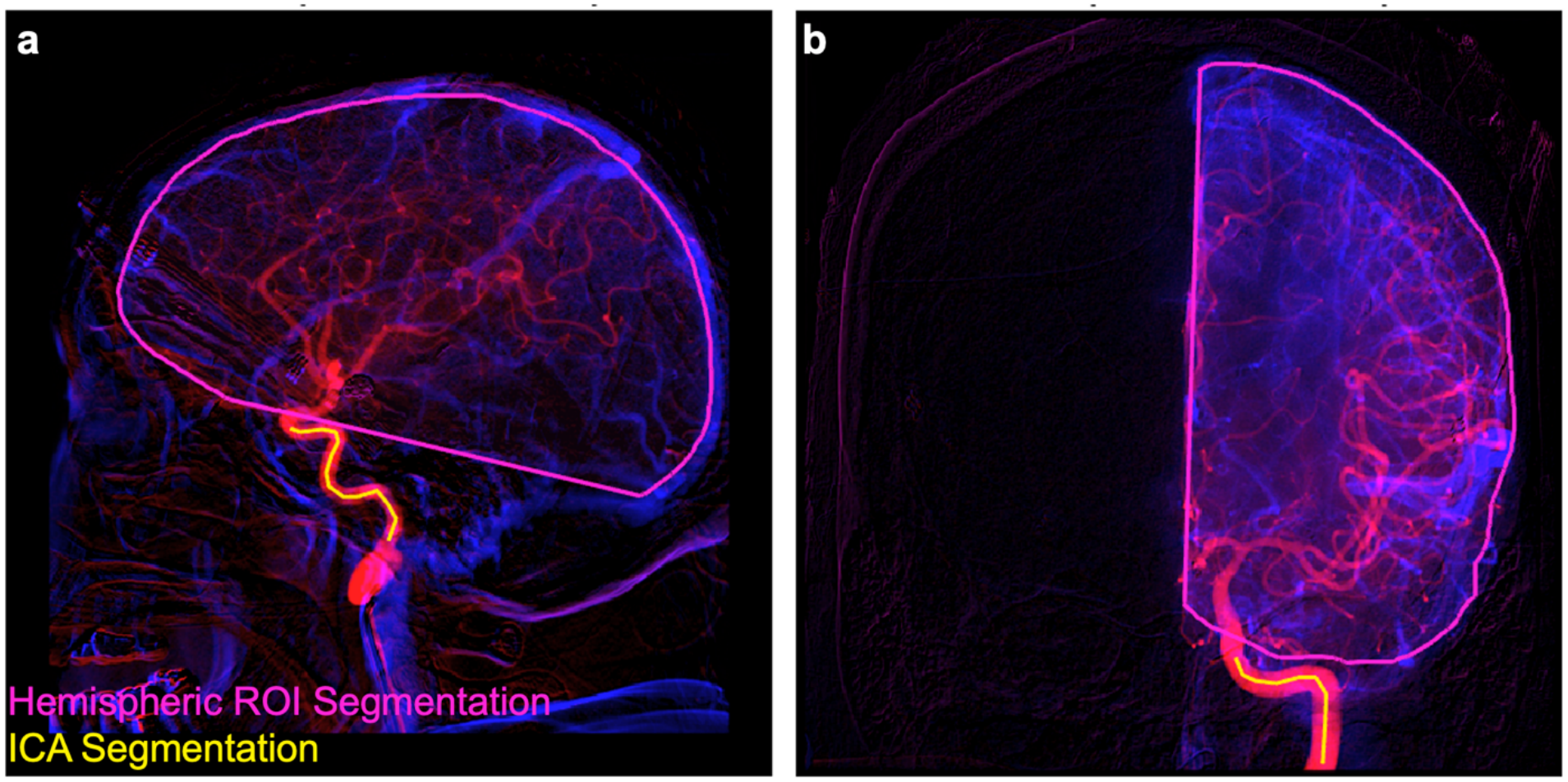
Manual anatomical reference annotations. Lateral (a) and AP (b) reference annotations on arterial/venous composites. Hemisphere contours define tissue ROIs. ICA centerlines, dilated by a radius-two disk on the 512 grid, define arterial segmentation targets.

Original frames and acquisition timing provided three hemisphere-model channels: arterial enhancement, late venous enhancement, and time-integrated enhancement. Enhancement was the positive difference between baseline and frame intensity. The arterial frame was selected from a smoothed global enhancement curve; late venous enhancement used the final three frames. Channels were independently percentile-scaled and resized to 256 × 256 (Fig. 3). Online Resource 1 details baseline estimation, frame selection, integration, and normalization.

**Figure 3.**
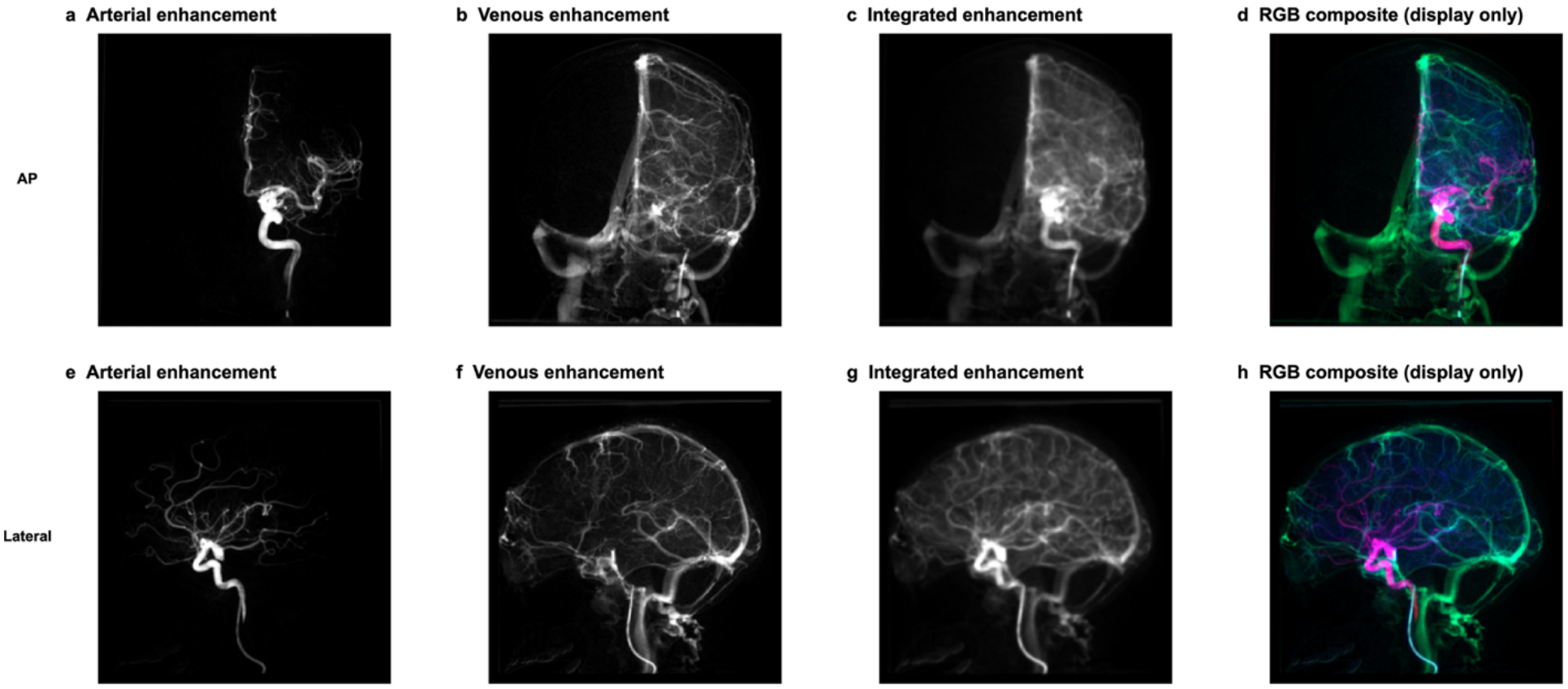
Hemisphere model input channels. AP (a–d) and lateral (e–h) examples share a patient and study day. Arterial (a, e), late venous (b, f), and integrated enhancement (c, g) form the normalized 256 × 256 inputs. RGB composites (d, h) display these channels in red, green, and blue. Grayscale channels share a 0–1 scale.

### Patient separated model development and testing

A master partition assigned internal patients to five folds, shared across projections and segmentation tasks. In outer round *k*, fold *k* provided testing, the next cyclic fold provided development validation, and the remaining three folds provided weight updates. Models were initialized afresh in each round. Every eligible internal run was evaluated once, in its patient’s outer test fold [15].

Separate candidates were trained with initial learning rates of 0.0001 and 0.001, without warm-up. Each candidate was trained for at most 120 epochs, with validation approximately once per epoch and early stopping after 25 consecutive validation checks without improvement in validation loss. The minimum-validation-loss checkpoint was restored. Learning rates were halved every 15 epochs according to a fixed schedule, independently of validation performance.

For each restored candidate, foreground-probability thresholds were evaluated from 0.5 to 0.9 for hemisphere segmentation and from 0.3 to 0.7 for ICA segmentation, in increments of 0.1. The candidate and threshold maximizing patient-mean validation Dice were selected, averaging runs within each patient before weighting patients equally. The candidate learning rates and threshold ranges were fixed development choices. Selected models and thresholds were applied unchanged to the corresponding test patients. Test predictions did not inform stopping, checkpoint restoration, or parameter selection [16].

For external evaluation, four fresh final models, one for each segmentation task and projection, were fitted on internal folds 1–4 and selected using fold 5 under the same procedure. External patients were excluded from both fitting and validation-based selection. Table S3 in Online Resource 1 details the training and selection protocol. Tables S4–S6 document the patient partitions and retained models.

### ResNet50 architecture and optimization

All four models used DeepLabv3+ with a ResNet50 encoder, combining deep residual feature extraction with multiscale contextual processing and boundary refinement [17–19]. This architecture was held fixed across AP and lateral projections and hemisphere and ICA segmentation tasks. Networks used an output stride of 16 and 256 × 256 × 3 inputs. Encoders were initialized with ImageNet-pretrained weights and fine-tuned jointly with newly initialized segmentation layers [20]. Table S3 in Online Resource 1 provides the complete training and selection protocol. Adam optimization used mini-batches of four, class-weighted loss, and L2 regularization [19]. Training augmentation paired image and label transforms: horizontal reflection, small rotations, translations, and independent axis scaling.

### Automatic hemisphere ROIs and ICA localization

ICA inputs comprised an automatically determined hemisphere ROI mask, arterial enhancement image, and time-integrated enhancement image, each resized to 256 x 256. Hemisphere ROI segmentation was trained using inner patient-separated cross-fitting so that each target patient was excluded from hemisphere ROI-model fitting and validation. For final validation, external testing, and deployment, hemispheric ROI were generated from the selected ResNet50 hemisphere model. Outer-test ICA inference used the corresponding patient-held-out ResNet50 hemisphere ROIs. Hemisphere probabilities were upsampled to 512 x 512, smoothed, thresholded, morphologically cleaned, and restricted to the largest connected component. The final hemisphere segmentation used its validation-selected cutoff (0.7 AP and 0.6 lateral).

### Projection routing and AIF selection

ICA probability maps were upsampled from 256 x 256 to 512 x 512 before connected-component cleanup, skeleton pruning, and graph-path centerline selection. Candidate AIF search ranges were selected by visual inspection to restrict sampling to anatomically suitable ICA segments. AIF candidate locations used AP centerline path fractions 0.40-0.60 and while lateral centerline fractions were 0.75-0.85. These values represent distance from the edge of the frame along the detected centerline of the ICA. Perpendicular intensity profiles supplied full-width-at-half-maximum estimates for cylindrical-width correction. Eligible weighted sampling regions were ranked by late-frame return toward baseline to reduce venous contamination.

### Perfusion calculation

Both the manual and automatic pipelines used the established perfusion routines [8, 11, 12]. Bolus arrival time (BAT) was the AIF’s maximum second derivative [21]. Block-circulant singular value decomposition used a relative threshold of 0.2 [22, 23]. Cerebral blood volume (CBV) values above the 70th percentile of nonzero values within each hemisphere were excluded to reduce vascular contamination [8]. Time to maximum residue (Tmax) was measured in the initial, non-extrapolated deconvolution [24–26]. Gamma-variate tails extended signals to 12 seconds after BAT when needed but longer measured series were not truncated [11]. Mean transit time (MTT) was CBV divided by cerebral blood flow (CBF) in the final output. Acquisition means used finite, strictly positive values within each workflow’s surviving-pixel mask. CBF and CBV remained relative measures without absolute calibration [27].

### Automatic versus manual perfusion

Eligible runs for manual and automated pipelines required at least 7 seconds of data after each respective BAT. The manual pipeline replaced learned localization and centerline-based AIF selection with the annotated hemisphere and AIF rectangle. Both pipelines were cylindrically corrected but each pipeline’s downstream calculations retained their own AIF, BAT, and surviving pixel mask.

### Segmentation evaluation

Endpoints were Dice, Jaccard, sensitivity, precision, mean symmetric boundary distance, and 95th-percentile Hausdorff distance (HD95) [28]. Distances used eight-connected boundaries on the 512 x 512 image. HD95 pooled bidirectional nearest-boundary distances. ICA predictions were compared with radius-two reference masks. Runs were averaged within patients, and 95% percentile confidence intervals resampled patients 2,000 times within cohort and projection.

### Biplane analysis

Independently processed AP and lateral runs were linked using adjudicated patient, study-day, and vessel identities. Both runs required at least 7 seconds after BAT [11].

### Statistical analysis

Manual comparisons weighted acquisitions equally and reported automatic-minus-manual bias, mean absolute error (MAE), and marginal Bland–Altman limits of agreement: mean difference ±1.96 times the sample standard deviation of paired differences [29]. Confidence intervals used 2,000 whole-patient bootstrap samples within cohort and projection, preserving repeated runs.

Segmentation summaries instead weighted patient-level averages equally.

Biplane comparisons reported mean absolute AP–lateral differences (MAE), weighting pairs equally. Confidence intervals for MAE used 2,000 whole-patient bootstrap samples within each cohort, preserving repeated pairs. Signed AP-minus-lateral differences were analyzed using intercept-only mixed-effects models with patient and patient-day-vessel random intercepts. The fixed intercept estimated bias, with 95% confidence intervals based on Kenward–Roger inference. Repeated-measures 95% limits of agreement were calculated as bias ±1.96 times the square root of the summed patient, patient-day-vessel, and residual variances. Models were fitted using restricted maximum likelihood. Median absolute differences and interquartile ranges provided complementary descriptive summaries. Scatter plots additionally showed mixed-effects AP-on-lateral regression.

A post hoc sensitivity analysis excluded pairs with AP MTT below 1 second or AP Tmax at least 10 seconds; the primary analysis retained these pairs. Processing and figure generation used MATLAB (R2025a, MathWorks, Natick, MA, USA). Biplane models used SAS PROC MIXED in SAS Studio online through SAS OnDemand for Academics.

## Results

### Eligible acquisitions and training behavior

Segmentation included 841 runs from 140 patients: 488 internal runs from 67 patients and 353 external runs from 73 patients. The external set comprised 182 AP runs from 64 patients and 171 lateral runs from 62 patients. Patients with both views contribute to both projection-specific counts. Fig. S1 and Table S6 in Online Resource 1 show training histories, retained checkpoints, and final model selections.

### External validation of hemisphere and ICA segmentation

External patient-mean hemisphere Dice was 0.959 (95% CI, 0.952–0.965) for AP and 0.965 (0.960–0.969) for lateral views; HD95 was 14.83 (95% CI, 12.82-17.11) and 15.62 (13.34 – 18.42) pixels. ICA Dice was 0.635 (0.617–0.650) and 0.558 (0.534–0.579), with HD95 of 13.65 (95% CI, 10.36-17.96) and 35.80 (30.18-42.48) pixels (Table 1; Figs. 4–5). Tables S7–S9 and Figs. S2–S3 in Online Resource 1 provide internal test results and all six metrics.

**Figure 4.**
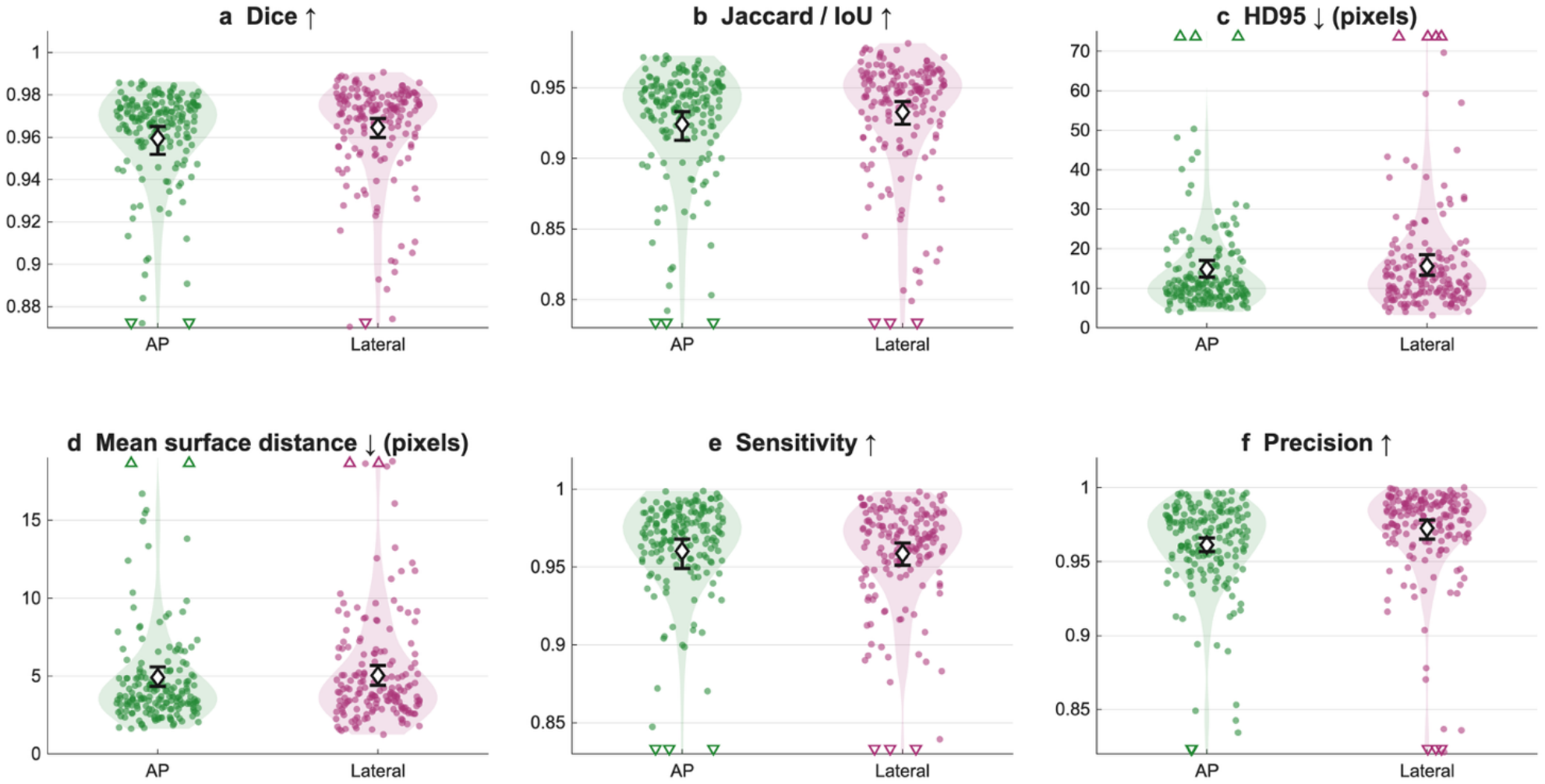
Hemisphere segmentation in the external validation cohort. Dice (a), Jaccard (b), HD95 (c), mean symmetric boundary distance (d), sensitivity (e), and precision (f) of the hemispheric segmentation model applied to the external cohort. Green AP and magenta lateral dots denote runs while shading shows distributions. Diamonds and whiskers show patient-mean scores and 95% patient-bootstrap intervals. Counts are 182 AP runs/64 patients and 171 lateral runs/62 patients. Triangles mark off-scale values.

**Figure 5.**
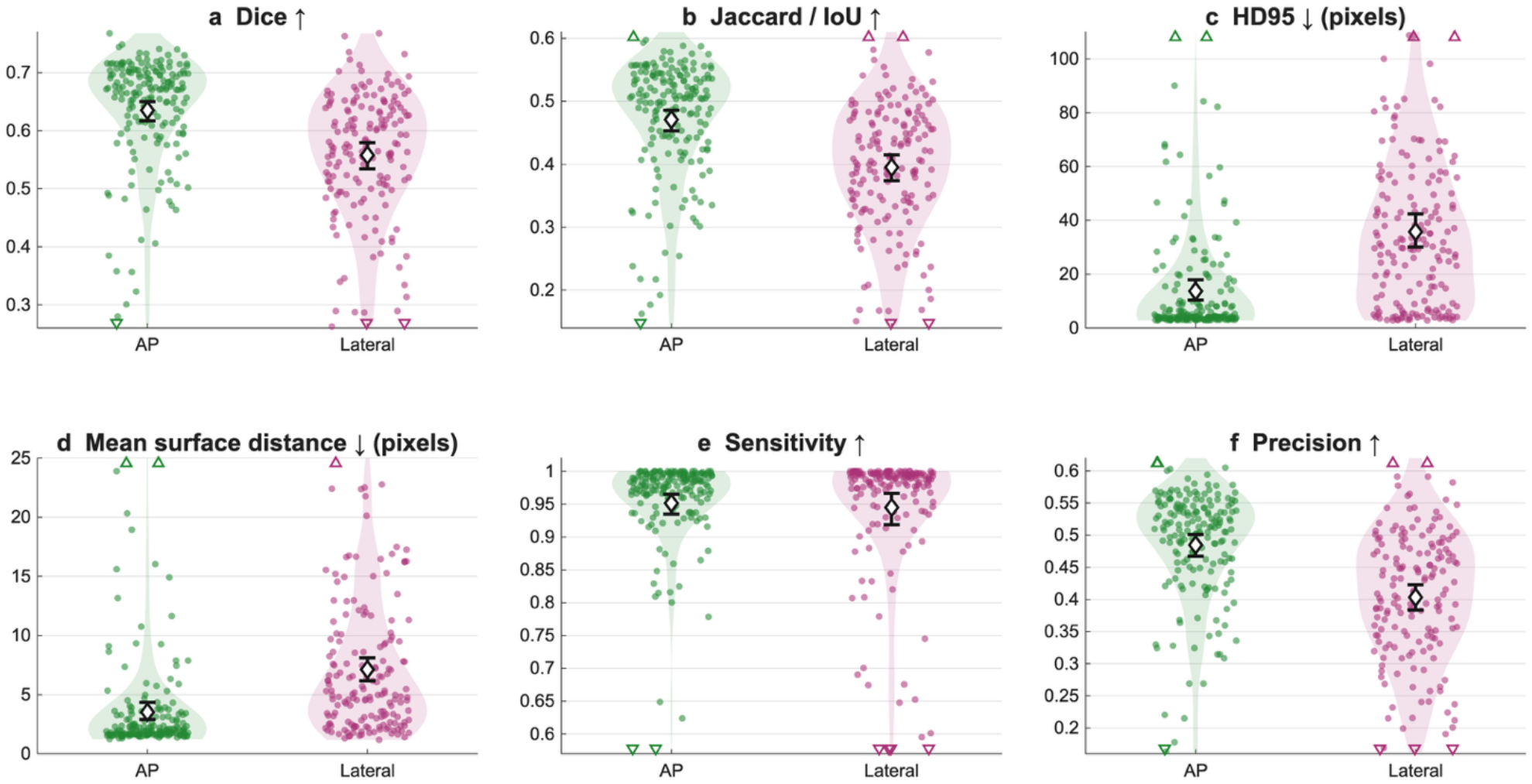
ICA segmentation in the external validation cohort. Panels, colors, sample sizes, and summaries follow Fig. 4. Dice (a), Jaccard (b), HD95 (c), mean symmetric boundary distance (d), sensitivity (e), and precision (f) are shown for ICA segmentation in the external cohort. Predictions are compared with manual ICA centerlines dilated by a radius-two disk. Metrics evaluate vessel localization before geometric centerline extraction and AIF selection.

**Table 1.**
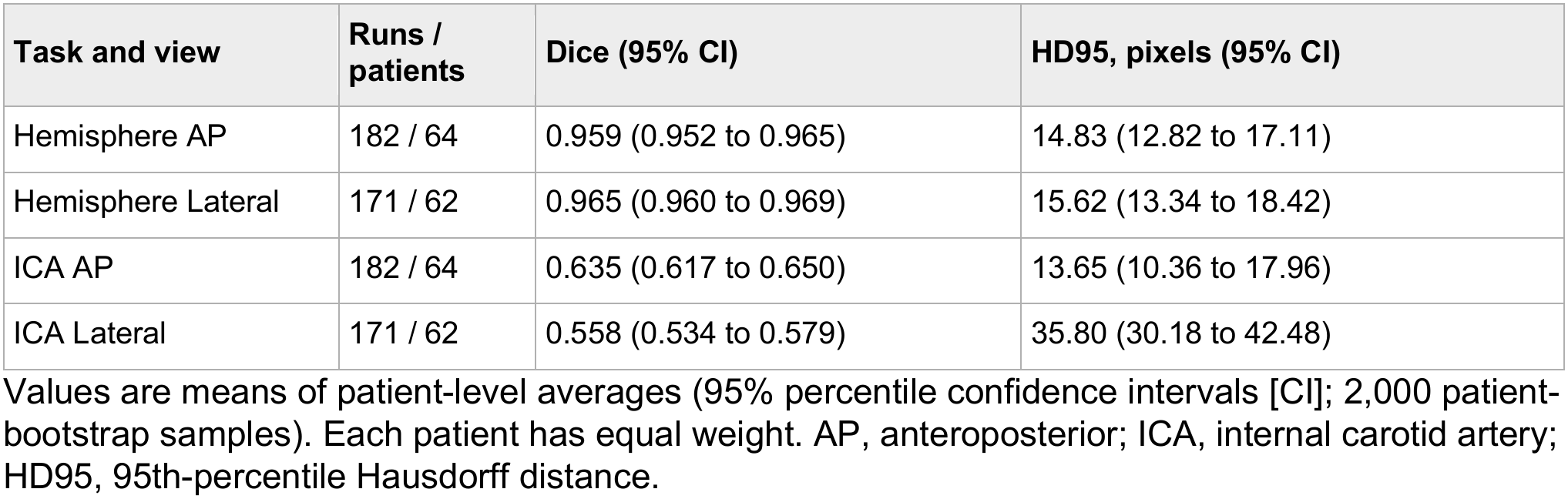
ResNet50 segmentation in the external validation cohort.

| **Task and view** | **Runs / patients** | **Dice (95% CI)** | **HD95, pixels (95% CI)** |
|---|---|---|---|
| Hemisphere AP | 182 / 64 | 0.959 (0.952 to 0.965) | 14.83 (12.82 to 17.11) |
| Hemisphere Lateral | 171 / 62 | 0.965 (0.960 to 0.969) | 15.62 (13.34 to 18.42) |
| ICA AP | 182 / 64 | 0.635 (0.617 to 0.650) | 13.65 (10.36 to 17.96) |
| ICA Lateral | 171 / 62 | 0.558 (0.534 to 0.579) | 35.80 (30.18 to 42.48) |

Values are means of patient-level averages (95% percentile confidence intervals [CI]; 2,000 patient-bootstrap samples). Each patient has equal weight. AP, anteroposterior; ICA, internal carotid artery; HD95, 95th-percentile Hausdorff distance.

### Automatic versus manual perfusion in the External validation cohort

The external cohort automatic versus manual pipeline comparison included 165 runs from 52 patients: 84 AP runs from 40 patients and 81 lateral runs from 44 patients. Figs. 6–7 illustrate close agreement; Table 2 and Fig. 8 summarize the cohort.

**Figure 6.**
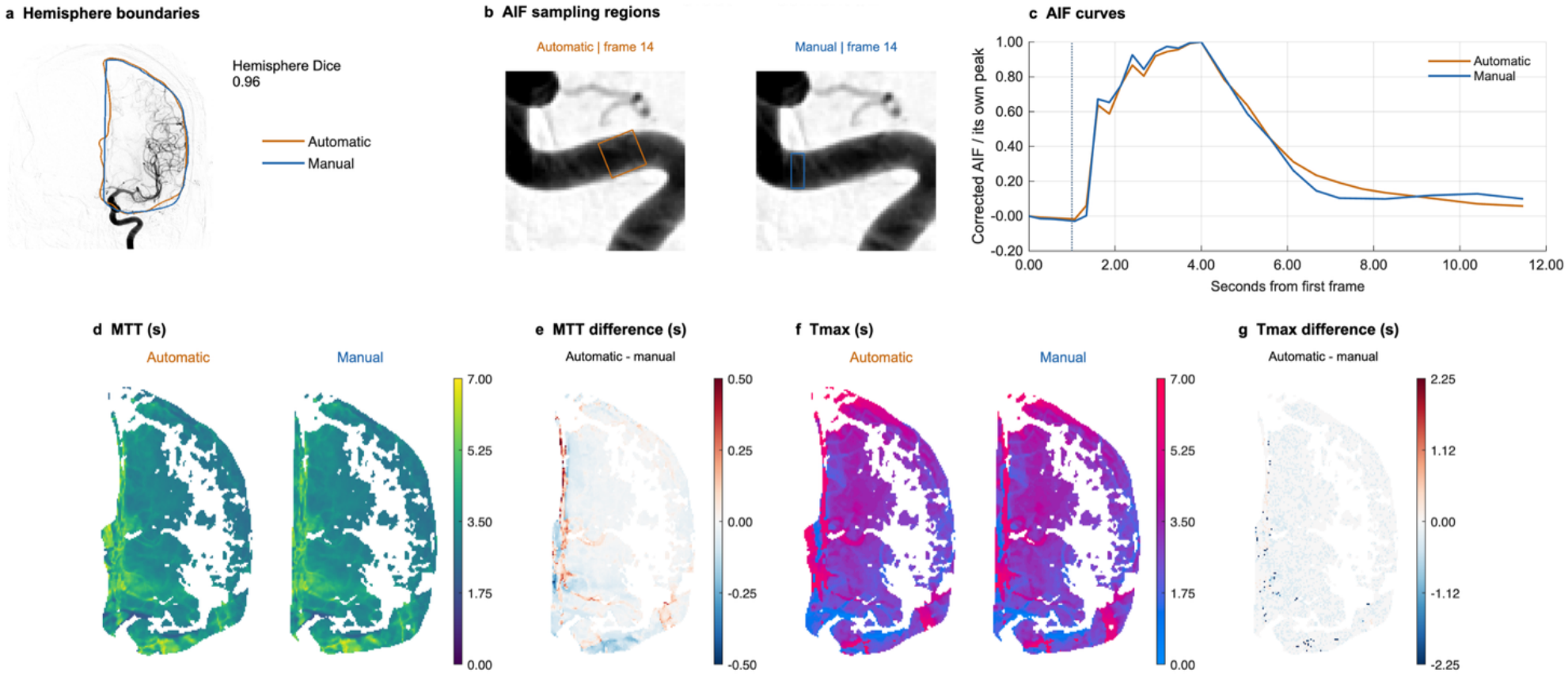
Representative AP agreement with manual-input perfusion. The automatic pipeline was applied to an external cohort example and compared with results obtained from manual processing. Hemisphere boundaries (a), solid AIF rectangles (b), and peak-normalized AIF curves (c); dotted vertical lines indicate bolus arrival time (BAT). Automatic (orange; left) and manual (blue; right) MTT (d) and Tmax (f) maps share scales. Difference maps (e, g) use common pixels. Hemisphere Dice measures anatomical overlap.

**Figure 7.**
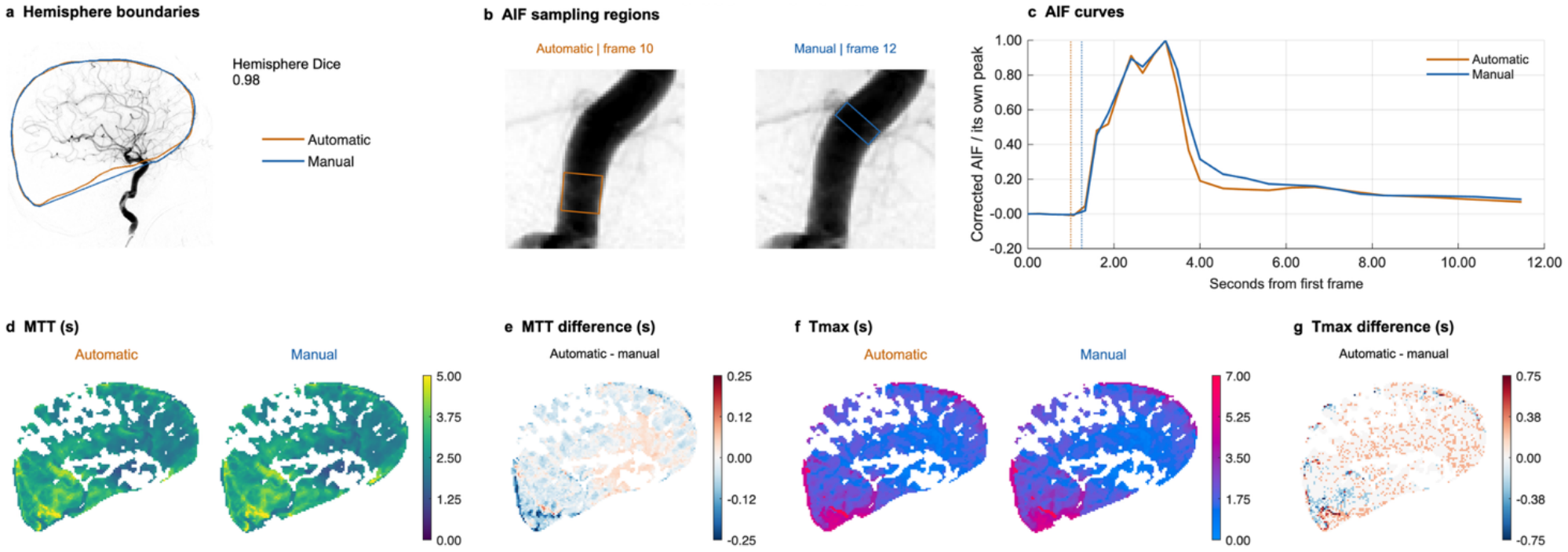
Representative lateral agreement with manual-input perfusion. Panels and colors follow Fig. 6 but comparing the automatic pipeline’s results from a lateral image with results obtained from manual processing. Hemisphere boundaries (a), solid AIF rectangles (b), and peak-normalized AIF curves (c); dotted vertical lines indicate bolus arrival time (BAT). Automatic (orange; left) and manual (blue; right) MTT (d) and Tmax (f) maps share scales. Difference maps (e, g) use common pixels. Hemisphere Dice measures anatomical overlap.

**Figure 8.**
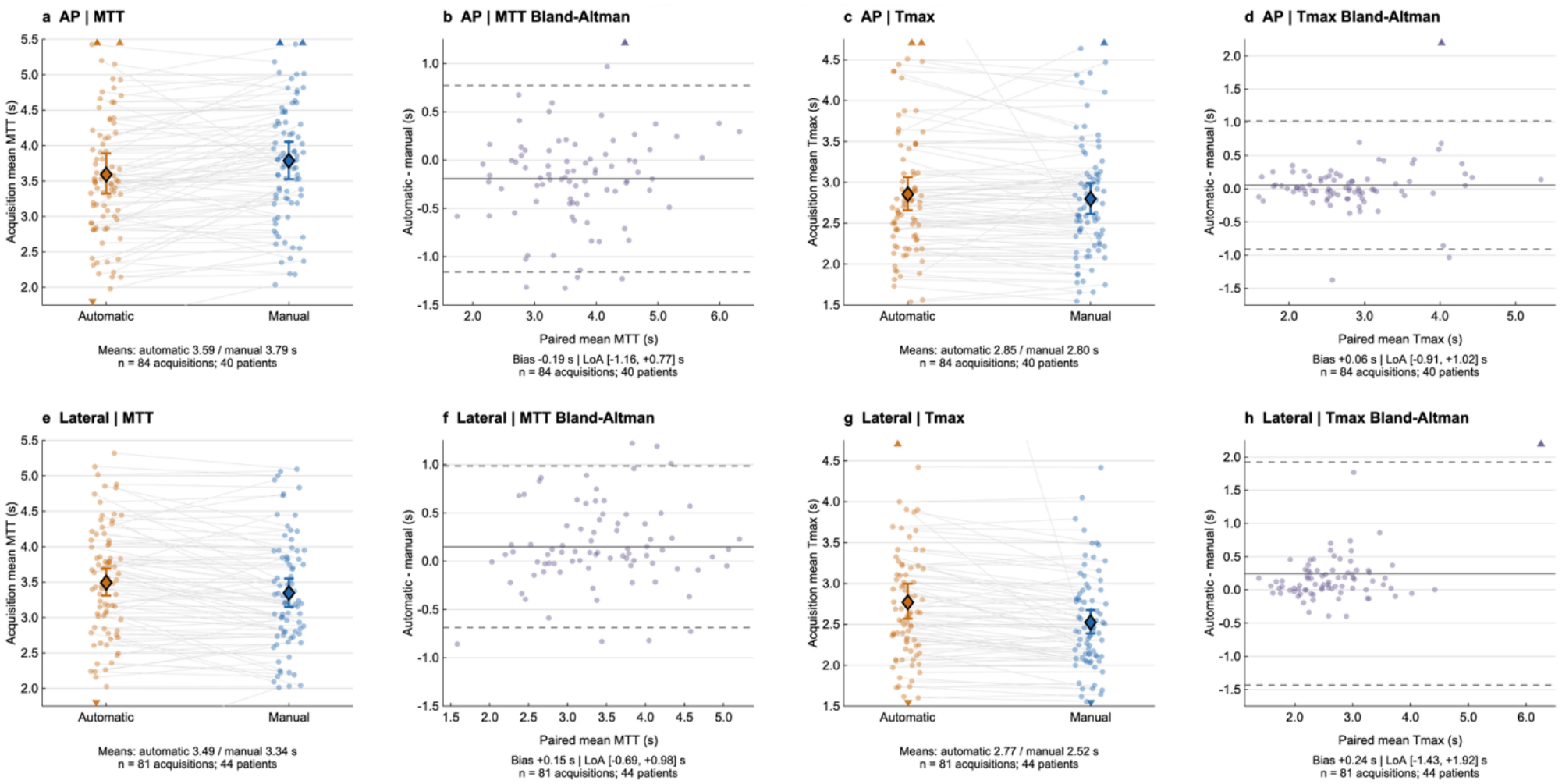
Automatic versus manual-input perfusion in the external validation cohort. AP (upper row, a-d) and lateral (lower row, e-h) acquisition means and Bland–Altman plots for MTT (first two columns, a, e) and Tmax (c, h). Orange/blue paired dots denote automatic/manual values; diamonds and whiskers show means and 95% whole-patient-bootstrap intervals. Bland–Altman differences are automatic minus manual; solid/dashed lines show bias/95% limits of agreement. Counts are 84 AP runs/40 patients and 81 lateral runs/44 patients, with 52 patients overall. Triangles mark off-scale observations.

**Table 2.**
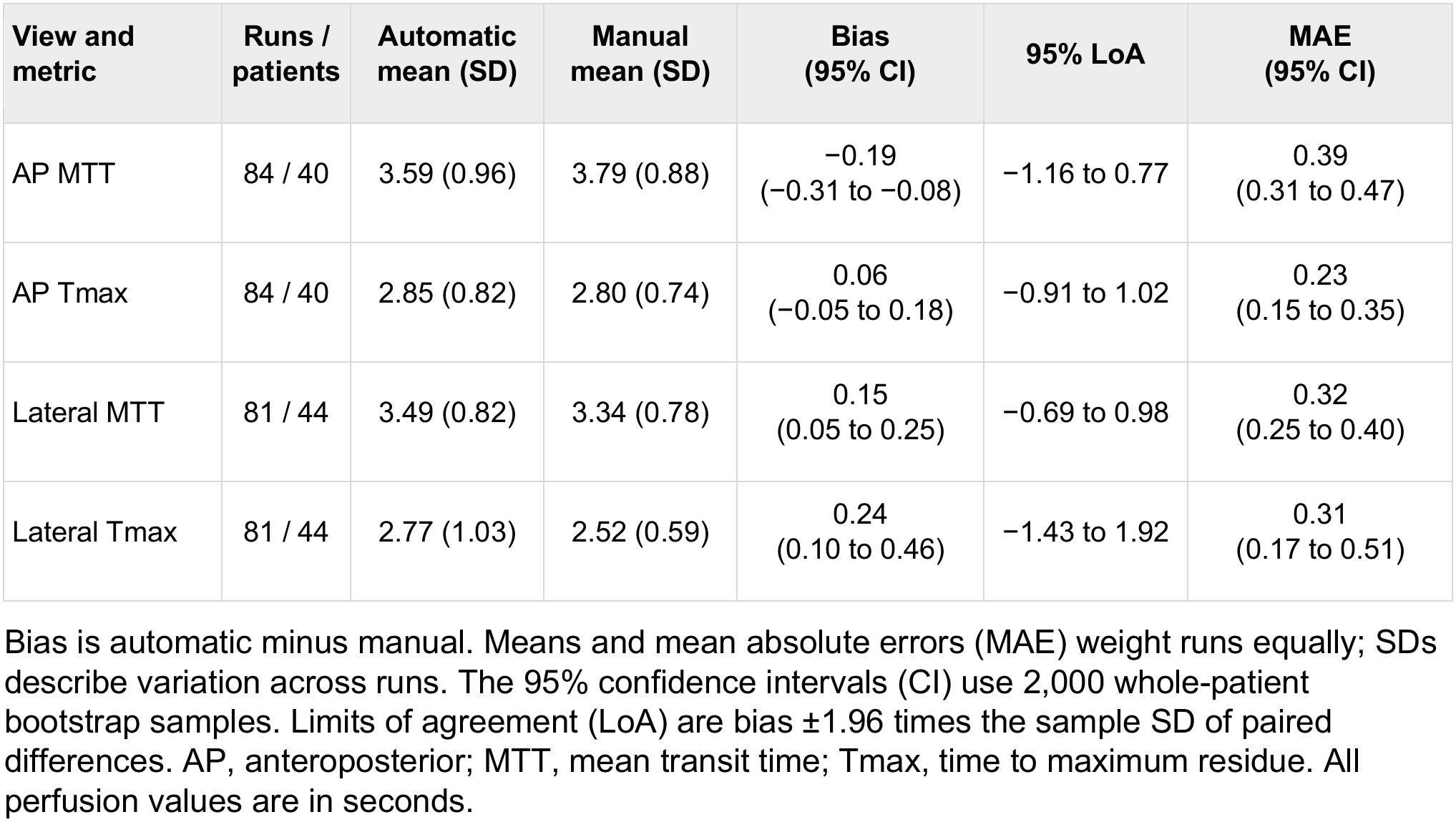
Automatic versus manual perfusion in the external validation cohort.

| **View and metric** | **Runs / patients** | **Automatic mean (SD)** | **Manual mean (SD)** | **Bias (95% CI)** | **95% LoA** | **MAE (95% CI)** |
|---|---|---|---|---|---|---|
| AP MTT | 84 / 40 | 3.59 (0.96) | 3.79 (0.88) | −0.19<br>(−0.31 to −0.08) | −1.16 to 0.77 | 0.39<br>(0.31 to 0.47) |
| AP Tmax | 84 / 40 | 2.85 (0.82) | 2.80 (0.74) | 0.06<br>(−0.05 to 0.18) | −0.91 to 1.02 | 0.23<br>(0.15 to 0.35) |
| Lateral MTT | 81 / 44 | 3.49 (0.82) | 3.34 (0.78) | 0.15<br>(0.05 to 0.25) | −0.69 to 0.98 | 0.32<br>(0.25 to 0.40) |
| Lateral Tmax | 81 / 44 | 2.77 (1.03) | 2.52 (0.59) | 0.24<br>(0.10 to 0.46) | −1.43 to 1.92 | 0.31<br>(0.17 to 0.51) |

Bias is automatic minus manual. Means and mean absolute errors (MAE) weight runs equally; SDs describe variation across runs. The 95% confidence intervals (CI) use 2,000 whole-patient bootstrap samples. Limits of agreement (LoA) are bias ±1.96 times the sample SD of paired differences. AP, anteroposterior; MTT, mean transit time; Tmax, time to maximum residue. All perfusion values are in seconds.

Mean (SD) AP MTT was 3.59 (0.96) seconds from the automatic pipeline and 3.79 (0.88) seconds from the manual pipeline. Bias was −0.19 seconds (95% CI, −0.31 to −0.08), with MAE 0.39 seconds (95% CI, 0.31-0.47). Lateral mean (SD) MTT was 3.49 (0.82) seconds from the automatic pipeline and 3.34 (0.78) seconds from the manual pipeline with bias 0.15 seconds (95% CI, 0.05-0.25) and MAE 0.32 seconds (95% CI, 0.25-0.40).

Mean (SD) AP Tmax was 2.85 (0.82) seconds from the automatic pipeline and 2.80 (0.74) seconds from the manual pipeline with bias 0.06 second (95% CI, −0.05 to 0.18) and MAE 0.23 seconds) 95% CI, 0.15-0.35). Lateral mean (SD) Tmax was 2.77 (1.03) seconds from the automatic pipeline and 2.52 (0.59) seconds from the manual pipeline, with bias 0.24 seconds (95% CI, 0.10-0.46) and MAE 0.31 seconds (95% CI, 0.17-0.51). Bias intervals excluded zero for AP MTT, lateral MTT, and lateral Tmax, indicating systematic differences between the pipelines. Online Resource 1 includes representative examples of larger disagreement between the automatic and manual pipelines (Figs. S4-S5) and summaries of both the internal and external cohorts (Table S10, Fig. S6).

### Biplane consistency in the External validation cohort

Biplane analysis included 220 external pairs from 61 patients and 187 patient-day-vessel groups. Mean (SD) AP and lateral MTT were 3.51 (0.95) and 3.58 (0.82) seconds; Tmax was 2.95 (1.40) and 2.79 (0.73) seconds. Median absolute differences were 0.33 seconds (IQR, 0.15-0.68) for MTT and 0.22 seconds (0.11-0.48) for Tmax. These summaries describe typical cross-projection differences with mean absolute errors reported in Table S11.

Repeated-measures AP-minus-lateral bias was −0.07 seconds for MTT (95% CI, −0.19 to 0.05), with limits of −1.38 to 1.24 seconds. Tmax bias 0.16 seconds (−0.08 to 0.40), with limits of agreement −2.65 to 2.97 seconds (Table 3, Fig. 9).

**Figure 9.**
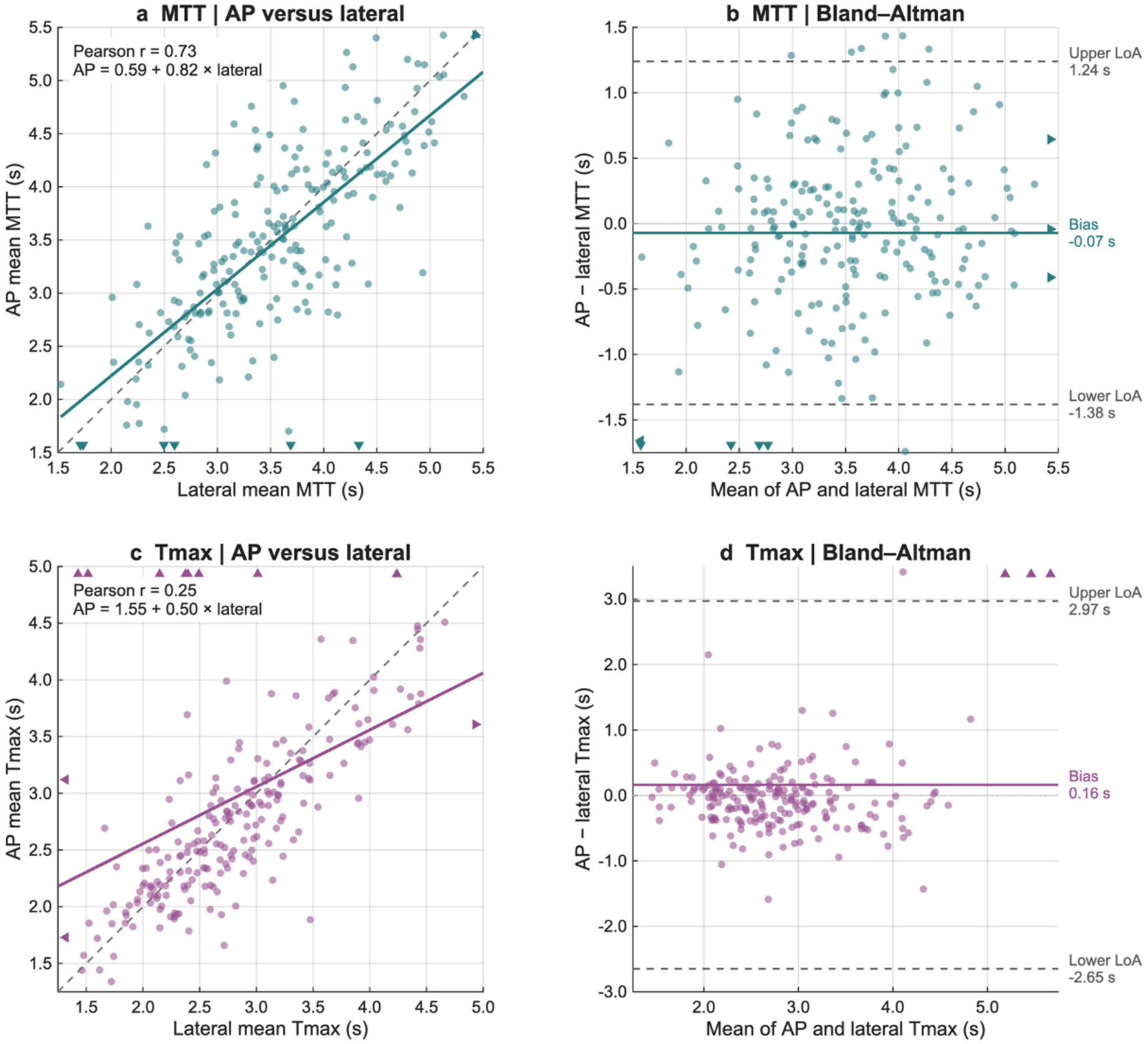
Biplane perfusion in the external validation cohort. MTT (a, b) and Tmax (c, d) scatter and Bland–Altman plots include all 220 pairs/61 patients. Scatter lines show identity (dashed) and mixed-effects regression (solid). AP-minus-lateral plots show mixed-model bias (solid) and repeated-measures 95% limits (dashed). Triangles indicate off-scale observations.

**Table 3.**
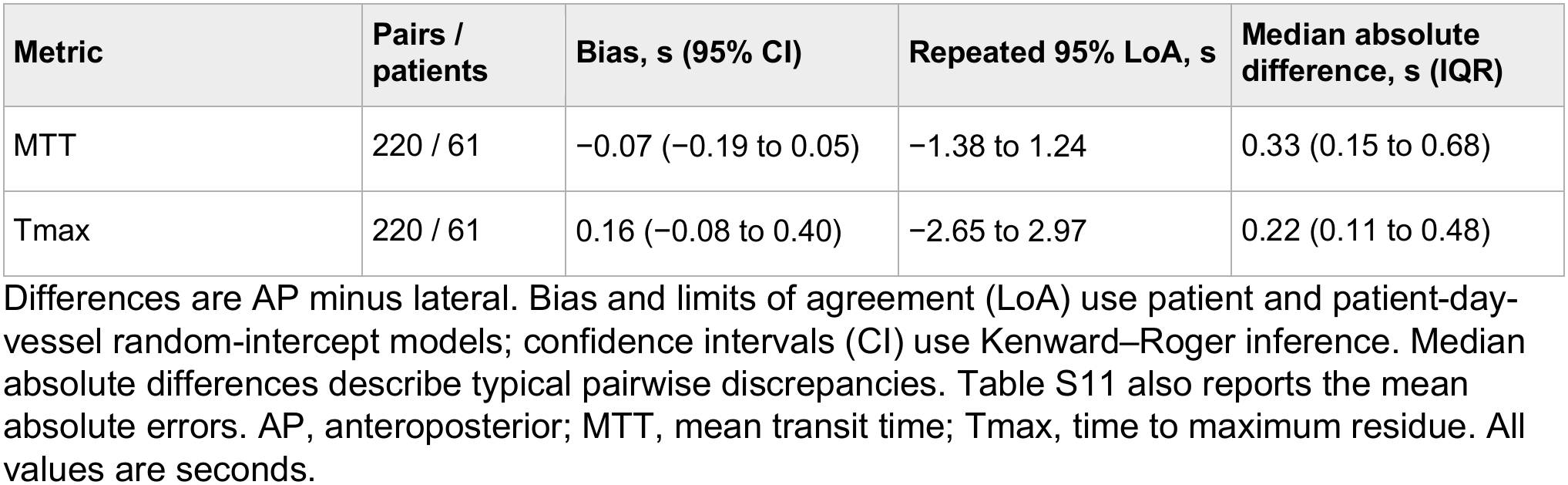
Automatic biplane agreement in the external validation cohort.

| Metric | Pairs / patients | Bias, s (95% CI) | Repeated 95% LoA, s | Median absolute difference, s (IQR) |
|---|---|---|---|---|
| MTT | 220 / 61 | −0.07 (−0.19 to 0.05) | −1.38 to 1.24 | 0.33 (0.15 to 0.68) |
| Tmax | 220 / 61 | 0.16 (−0.08 to 0.40) | −2.65 to 2.97 | 0.22 (0.11 to 0.48) |

Differences are AP minus lateral. Bias and limits of agreement (LoA) use patient and patient-day-vessel random-intercept models; confidence intervals (CI) use Kenward–Roger inference. Median absolute differences describe typical pairwise discrepancies. Table S11 also reports the mean absolute errors. AP, anteroposterior; MTT, mean transit time; Tmax, time to maximum residue. All values are seconds.

A post hoc review identified four biplane pairs with extreme AP perfusion estimates (MTT below 1 second or Tmax at least 10 seconds), prompting an exploratory sensitivity analysis to assess their influence on agreement. Excluding these pairs left 216 pairs from 60 patients. MTT bias was −0.03 seconds (95% CI, −0.13 to 0.07), with limits of agreement of −1.18 to 1.13 seconds; Tmax bias was 0.01 seconds (95% CI, −0.11 to 0.12), with limits of agreement of −1.56 to 1.57 seconds. These narrower limits illustrate the influence of extreme AP estimates. Because the exclusion criteria were defined post hoc, all 220 eligible pairs were retained in the primary analysis. Online Resource 1 reports the descriptive and repeated-measures comparisons (Tables S11–S12).

## Discussion

Digital subtraction angiography is traditionally interpreted as a measure of cerebrovascular anatomy, yet contrast passage also contains information about tissue perfusion. Here, we demonstrate a fully automated pipeline for extracting this information without manual anatomical localization. In the held-out external cohort, mean absolute differences from manual-input processing were 0.32–0.39 seconds for MTT and 0.23–0.31 seconds for Tmax. Nevertheless, automated processing produced higher lateral MTT and Tmax, with biases of 0.15 seconds (95% CI, 0.05–0.25) and 0.24 seconds (0.10–0.46), respectively, and lower AP MTT, with a bias of −0.19 seconds (−0.31 to −0.08). These intervals exclude zero, indicating systematic differences between the pipelines. Independently processed AP and lateral projections also showed small average biases, although variability between individual measurements limits their interchangeability.

The clinical significance of these differences cannot be determined from agreement alone, as there are not yet clinically meaningful thresholds for angiographic MTT or Tmax. Furthermore, the manually processed pipeline served as a methodological comparator, not an independent physiological reference standard. Because both pipelines share the same downstream perfusion calculations, agreement does not establish the accuracy of either method. The automated pipeline may nonetheless provide a practical means of processing large cohorts consistently, enabling clinical research into associations between angiographic perfusion, treatment response, and patient outcomes. Such studies are needed to determine which perfusion measurements and changes are clinically informative.

The clinical significance of this approach lies in the opportunity to measure physiological responses to cerebrovascular interventions rather than relying exclusively on changes in vascular anatomy. This distinction is particularly relevant to endovascular rescue therapy (ERT) for vasospasm following aneurysmal subarachnoid hemorrhage (aSAH), where angiographic narrowing, delayed cerebral infarction, and functional outcome represent related but distinct endpoints [1–3]. In our previous single-center retrospective investigations, ERT was associated with improvements in angiographic perfusion metrics and subsequent time within the cerebral autoregulatory range [8, 11, 12]. However, establishing the clinical relevance of these physiological measurements requires substantially larger and more diverse patient populations. By enabling reproducible analysis across institutions, the present workflow provides an opportunity to investigate whether changes in angiographic perfusion following ERT reflect meaningful physiological improvement, predict subsequent cerebral infarction, or provide information beyond conventional assessments of vessel caliber.

Our findings build on a growing body of work demonstrating the feasibility of quantitative perfusion analysis from planar angiography. Previous investigators have developed automated arterial input function (AIF) extraction and deconvolution-based perfusion mapping with perfDSA, as well as approaches for rapid perfusion processing during angiographic acquisition [10, 13]. The present study extends these efforts by integrating projection-specific hemisphere and internal carotid artery localization within a unified workflow. We evaluated performance in a held-out institutional cohort and assessed agreement between independently processed biplane projections. These complementary evaluations address reproducibility across institutions and consistency across angiographic acquisition geometries, both relevant to broader adoption.

An important methodological consideration is that accurate identification of an AIF does not necessarily require anatomically perfect segmentation of the internal carotid artery. The physiological information used for deconvolution resides in the sampled arterial contrast-time curve rather than in the geometric fidelity of the arterial segmentation itself. Consistent with this distinction, previous work with perfDSA demonstrated that imperfect automated ICA segmentations could nevertheless produce arterial curves closely resembling those obtained from manual annotations [10, 13]. Conversely, investigations of CT perfusion have demonstrated that AIF selection can materially influence downstream perfusion estimates [30].These observations suggest that segmentation overlap alone is an incomplete measure of successful automated arterial localization.

Our biplane analysis further highlights the distinction between reproducible average measurements and agreement at the individual acquisition level. Although AP and lateral projections demonstrated small median absolute differences and limited average bias, agreement was stronger for MTT than for Tmax. In particular, the primary Tmax analysis revealed substantial tails in the distribution of paired differences. These findings are consistent with the fundamentally different tissue information represented by planar projections acquired from different geometries and emphasize the importance of projection-specific interpretation.

Several considerations remain important before this approach can be applied broadly. Although patients in the external validation cohort were excluded from model development and selection, agreement with manual-input processing does not independently establish physiological accuracy or generalizability across all acquisition protocols. Furthermore, successful automation does not eliminate the need for quality assurance. As with clinically implemented CT perfusion software, we envision an automated workflow accompanied by targeted review of quality-control outputs, including arterial sampling location, AIF morphology, hemisphere segmentation, and resulting perfusion maps [31, 32].

Ultimately, the most important unanswered question is not whether angiography can provide quantitative perfusion measurements, but whether those measurements can improve the assessment and management of cerebrovascular disease. Although observational studies have investigated axial perfusion imaging in aSAH [33–35], the optimal perfusion parameters and thresholds for guiding treatment remain uncertain [3]. Furthermore, the relationship between planar angiographic perfusion and established axial perfusion modalities has not been systematically characterized in large, diverse cohorts. Future investigations should therefore prioritize cross-modal validation and prospective evaluation of associations with clinically meaningful outcomes. By making quantitative perfusion analysis feasible across large angiographic datasets, this approach creates an opportunity to investigate not only how cerebrovascular interventions change arterial anatomy, but whether they restore the perfusion of the tissue those arteries supply.

## Conclusion

The automated perfusion pipeline showed favorable agreement with manual processing in the held-out external validation cohort. Biplane views had small typical differences and average bias. These results support automated angiographic perfusion research with user quality control review.

## Statements and Declarations

### Ethics approval and consent to participate

Institutional review boards at Massachusetts General Hospital and Yale New Haven Hospital approved this retrospective study and waived informed consent.

### Authors’ contributions

All authors conceived and designed the study. KL and LPT prepared material, collected data, and performed analyses. KL drafted the manuscript; all authors revised and approved it.

### Data avxailability

Data supporting this study are available from the corresponding author on reasonable request, subject to institutional approval.

### Code availability

Code supporting this study is available from the corresponding author on reasonable request, subject to institutional approval.

