## SupplementaryMaterial for "Automatic cerebral angiographic perfusion analysis"

**Online Resource 1 Supplementary methods and results**

Abbreviations: AIF, arterial input function; AP, anteroposterior; BAT, bolus arrival time; CI, confidence interval; DICOM, Digital Imaging and Communications in Medicine; HD95, 95th-percentile Hausdorff distance; ICA, internal carotid artery; LoA, limits of agreement; MTT, mean transit time; QC, quality control; REML, restricted maximum likelihood; ROI, region of interest; Tmax, time to maximum of the residue function.

**Supplementary Methods**
**Supplementary Tables S1-S12**
**Supplementary Figures S1-S6**

### Supplementary methods

#### Input preparation
Baseline intensity was the mean of the first approximately 8% of frames, constrained to two to four frames. Positive enhancement was baseline intensity minus frame intensity, floored at zero. The arterial frame was selected from a temporally smoothed global 99.7th-percentile enhancement curve, but the arterial input channel used unsmoothed enhancement at that frame. The venous channel was baseline minus the mean of the final three frames. Integrated enhancement used trapezoidal time integration of temporally smoothed enhancement, followed by Gaussian smoothing with sigma two pixels. Each channel was independently clipped and scaled at the first and 99.7th percentiles and bilinearly resized to 256 × 256.

#### Model fitting and selection
Adam used moment decay factors 0.9 and 0.999, epsilon $1 \times 10^{-8}$, and L2 regularization 0.0001. Background class weight was one and foreground weight was the training-mask background/foreground pixel ratio, capped at 50. MATLAB constructor learning-rate factors were retained, including factor 10 in added trainable convolutions and zero in fixed upsampling layers. Independent candidates started at learning rates 0.0001 and 0.001, with no warm-up. A fixed schedule multiplied the base rate by 0.5 every 15 epochs. Training allowed at most 120 epochs. Validation occurred approximately once per epoch, and training stopped after 25 consecutive validation checks without improvement in loss. The minimum-validation-loss checkpoint was restored before evaluating candidate cutoffs (hemisphere, 0.5–0.9; ICA, 0.3–0.7; increments of 0.1). The candidate and cutoff maximizing patient-mean validation Dice were applied unchanged to test patients. Augmentation paired images and masks using random horizontal reflection, rotations from −8° to +8°, translations from −10 to +10 pixels, and independent horizontal/vertical scaling from 0.92 to 1.08. Validation and test images were not augmented. No additional dropout was introduced.
The shared master assignment retains a patient's fold across views. Across outer rounds, the training/validation/test fold schedules are 3,4,5/2/1; 1,4,5/3/2; 1,2,5/4/3; 1,2,3/5/4; and 2,3,4/1/5. A patient can train models in other rounds but contributes test predictions only when its own fold is held out so that roles never overlapped within a round.

#### Hemisphere ROIs and cleanup
Hemisphere ROIs supplied to the ICA training routines used inner patient-separated cross-fitting so that the target patient was excluded. Inner validation comprised 15% of the remaining training patients, rounded upward. This preparatory ROI generation was distinct from ICA-model fitting. Both segmentation networks received 256 × 256 × 3 inputs and produced 256 × 256 foreground-probability maps, bilinearly upsampled to 512 × 512 for cleanup. Hemisphere probabilities underwent Gaussian smoothing (sigma 2.5 pixels; 11-pixel filter), thresholding, closing with a radius-two disk, opening with a radius-one disk, hole filling, removal of eight-connected components smaller than 200 pixels, and retention of the largest component. Final hemisphere segmentation ROIs used validation-selected cutoffs (0.7 AP and 0.6 lateral for the final models).

#### AIF sampling and comparison eligibility
Automatic AIF candidates spanned centerline fractions 0.40–0.60 in steps of 0.05 for AP and 0.75–0.85 in steps of 0.01 for lateral views (default lateral fraction 0.80). Perpendicular full-width-at-half-maximum profiles informed cylindrical-width correction. The manual rectangle's long axis crossed the vessel and its short axis followed the vessel. Both workflows retained their own AIF, bolus-arrival estimate, and surviving tissue mask. The external primary comparison contained 165 runs/52 patients for both metrics.

#### Biplane analysis
Primary biplane results use all 220 eligible external pairs. Intercept-only REML mixed models contain random intercepts for patient and patient-day-vessel; fixed-effect intervals use Kenward–

Roger degrees of freedom. Limits of agreement equal bias ±1.96 times the square root of summed variance components. Table S12 shows the repeated-measures analysis and the post hoc exclusion of AP MTT below 1 second or AP Tmax at least 10 seconds.

**Supplementary Table S1. Cohort demographics**

| Cohort | n | Male sex, n (%) | Age, years, mean (SD) | Hunt–Hess grade, median [IQR] | Modified Fisher grade, median [IQR] |
|---|---|---|---|---|---|
| Internal validation | 67 | 21 (31.3) | 57.6 (14.1) | 3 [2–5] | 4 [3–4] |
| External validation | 78 | 22 (28.2) | 56.2 (13.4) | 3 [2–4] | 4 [4–4] |
| All patients | 145 | 43 (29.7) | 56.8 (13.7) | 3 [2–4] | 4 [4–4] |

Values are number (percentage), mean (standard deviation [SD]), or median [25th–75th percentile; interquartile range (IQR)]. Cohort characteristics are presented descriptively.

**Supplementary Table S2. Acquisition characteristics of the segmentation cohorts**

| Characteristic | Internal AP | Internal lateral | External AP | External lateral |
|---|---|---|---|---|
| Eligible acquisitions, n | 232 | 256 | 182 | 171 |
| Patients, n | 59 | 60 | 64 | 62 |
| Frames per acquisition | 26.0 [23.0, 30.0] | 25.0 [23.0, 29.0] | 31.0 [25.0, 33.8] | 31.0 [25.0, 34.0] |
| First-to-last-frame duration, s | 9.07 [8.00, 10.31] | 8.78 [7.52, 10.50] | 8.27 [7.50, 9.50] | 8.27 [7.50, 9.50] |
| Primary positioner angle, degrees | 0.1 [-2.0, 3.9] | -90.0 [-91.1, -87.0] | -0.1 [-8.8, 5.8] | -89.9 [-97.3, -84.2] |
| Secondary positioner angle, degrees | 0.9 [-5.1, 8.2] | 0.3 [-0.1, 4.3] | 18.8 [11.4, 25.5] | 0.1 [-1.3, 3.8] |
| Source matrix, pixels (n) | 1024 x 1024 (2); 1152 x 1152 (2); 1440 x 1440 (59); 512 x 512 (168); 928 x 928 (1) | 1024 x 1024 (2); 1440 x 1440 (8); 1856 x 1856 (60); 512 x 512 (186) | 1024 x 1024 (182) | 1024 x 1024 (171) |

Values are median [25th, 75th percentile] unless indicated. AP, anteroposterior. Duration is first-to-final-frame elapsed time; angles are raw DICOM positioner values.

#### Supplementary Table S3. Training and selection protocol

| Item | Implemented setting |
|---|---|
| Network and initialization | DeepLabv3+; ImageNet-initialized ResNet50 encoder with newly initialized segmentation layers |
| Training mode | Joint fine-tuning of trainable encoder and segmentation layers |
| Input and output stride | 256 × 256 × 3; stride 16 |
| Hemisphere channels | Arterial enhancement, late venous enhancement, time-integrated enhancement |
| ICA channels | Automatic hemisphere ROI, arterial enhancement, time-integrated enhancement |
| Reference ICA target | Manual centerline dilated by radius-two disk on the 512-pixel grid |
| Partition | Five patient-level folds shared by both projections/tasks |
| Outer train validation test roles | Three fitting folds; next cyclic fold for validation; one test fold |
| Final external workflow | Internal folds 1–4 for fitting; internal fold 5 for validation; external patients for testing |
| Candidate learning rates | 0.0001 and 0.001 |
| Optimizer | Adam; beta1 0.9; beta2 0.999; epsilon $1 \times 10^{-8}$ |
| Batch size and shuffling | Four; shuffled each epoch |
| Learning-rate schedule | No warmup used. Fixed piecewise schedule multiplying the base rate by 0.5 every 15 epochs |
| Regularization | L2 0.0001 |
| Class weights | Background 1; foreground training background/foreground pixel ratio, capped at 50 |
| Layer learning-rate factors | Constructor factors retained; added trainable convolution factors include 10; fixed upsampling layers retain 0 |
| Training duration | Maximum 120 epochs; patience 25 epochs |
| Retained checkpoint | Best validation-loss network, not the final recorded epoch |
| Training augmentation | Random horizontal reflection; rotation −8° to +8°; x/y translation −10 to +10 pixels; independent x/y scaling 0.92–1.08 |
| Hemisphere candidate thresholds | 0.5, 0.6, 0.7, 0.8, 0.9 |
| ICA candidate thresholds | 0.3, 0.4, 0.5, 0.6, 0.7 |
| Candidate and threshold selection | Maximum patient-mean validation Dice across restored candidates and cutoffs; stable first-candidate rule for ties; selection fixed before testing |
| Hemisphere ROIs for ICA training | Inner cross-fitted hemisphere predictions; target patient excluded from guide fitting and validation |
| Inner validation and ICA input ROI | 15% of remaining training patients, rounded upward |
| Uncertainty | 2,000 patient-level bootstrap resamples within cohort and projection |

**Supplementary Table S4. Patient separated outer-fold counts**

| View | Test fold | Training runs / patients | Validation runs / patients | Test runs / patients |
|---|---|---|---|---|
| AP | 1 | 142 / 37 | 44 / 11 | 46 / 11 |
| AP | 2 | 140 / 36 | 48 / 12 | 44 / 11 |
| AP | 3 | 137 / 34 | 47 / 13 | 48 / 12 |
| AP | 4 | 138 / 34 | 47 / 12 | 47 / 13 |
| AP | 5 | 139 / 36 | 46 / 11 | 47 / 12 |
| Lateral | 1 | 154 / 38 | 48 / 11 | 54 / 11 |
| Lateral | 2 | 157 / 36 | 51 / 13 | 48 / 11 |
| Lateral | 3 | 153 / 34 | 52 / 13 | 51 / 13 |
| Lateral | 4 | 153 / 35 | 51 / 12 | 52 / 13 |
| Lateral | 5 | 151 / 37 | 54 / 11 | 51 / 12 |

Counts are unaugmented acquisitions and distinct patients within each projection and role. Internal AP total: 232 runs/59 patients. Internal lateral total: 256 runs/60 patients.

**Supplementary Table S5. Final model fitting and external testing**

| View | Internal training runs / patients | Internal validation runs / patients | External runs / patients |
|---|---|---|---|
| AP | 185 / 47 | 47 / 12 | 182 / 64 |
| Lateral | 205 / 48 | 51 / 12 | 171 / 62 |

The final models used internal folds 1–4 for fitting and fold 5 for validation. These validation observations were used for model selection and are not an independent test set. The external validation set contains 73 distinct patients.

**Supplementary Table S6. Retained ResNet50 checkpoints**

| Task and projection | Initial rate | Threshold | Retained epoch | Last epoch | Best validation loss |
|---|---|---|---|---|---|
| AP hemisphere | 0.001 | 0.7 | 43 | 68 | 0.04559 |
| Lateral hemisphere | 0.0001 | 0.6 | 14 | 39 | 0.05341 |
| AP ICA | 0.001 | 0.7 | 23 | 48 | 0.01825 |
| Lateral ICA | 0.0001 | 0.7 | 21 | 46 | 0.02258 |

Selected epochs identify the minimum recorded validation loss. Initial learning rates precede scheduled halving and thresholds are foreground-probability cutoffs selected by patient-mean validation Dice. Fig. S1 shows the corresponding training histories.

**Supplementary Table S7. ResNet50 segmentation overlap**

| Cohort and task | View | Runs / patients | Dice (95% CI) | Jaccard (95% CI) |
|---|---|---|---|---|
| Internal validation Hemisphere | AP | 232 / 59 | 0.966 (0.963 to 0.969) | 0.935 (0.930 to 0.940) |
| Internal validation Hemisphere | Lateral | 256 / 60 | 0.971 (0.969 to 0.973) | 0.945 (0.941 to 0.948) |
| Internal validation ICA | AP | 232 / 59 | 0.635 (0.617 to 0.652) | 0.471 (0.454 to 0.488) |
| Internal validation ICA | Lateral | 256 / 60 | 0.605 (0.584 to 0.626) | 0.443 (0.423 to 0.464) |
| External validation Hemisphere | AP | 182 / 64 | 0.959 (0.952 to 0.965) | 0.924 (0.913 to 0.933) |
| External validation Hemisphere | Lateral | 171 / 62 | 0.965 (0.960 to 0.969) | 0.933 (0.924 to 0.940) |
| External validation ICA | AP | 182 / 64 | 0.635 (0.617 to 0.650) | 0.471 (0.453 to 0.486) |
| External validation ICA | Lateral | 171 / 62 | 0.558 (0.534 to 0.579) | 0.395 (0.374 to 0.415) |

Scores are patient-level means with runs averaged within each patient before equal weighting of patients. Parentheses give 95% percentile confidence intervals from 2,000 patient-bootstrap samples. All listed runs contribute finite scores and no empty predictions occurred. Figs. S2 and S3 show the individual observations and population summaries.

**Supplementary Table S8. ResNet50 segmentation boundary distances**

| Cohort and task | View | Runs / patients | HD95, pixels (95% CI) | Mean boundary distance, pixels (95% CI) |
|---|---|---|---|---|
| Internal validation Hemisphere | AP | 232 / 59 | 11.09 (10.09 to 12.21) | 4.03 (3.71 to 4.37) |
| Internal validation Hemisphere | Lateral | 256 / 60 | 12.19 (10.99 to 13.55) | 4.28 (3.93 to 4.68) |
| Internal validation ICA | AP | 232 / 59 | 16.77 (11.64 to 22.79) | 4.05 (3.23 to 4.96) |
| Internal validation ICA | Lateral | 256 / 60 | 31.78 (26.20 to 37.53) | 6.76 (5.33 to 8.47) |
| External validation Hemisphere | AP | 182 / 64 | 14.83 (12.82 to 17.11) | 4.90 (4.33 to 5.58) |
| External validation Hemisphere | Lateral | 171 / 62 | 15.62 (13.34 to 18.42) | 5.02 (4.41 to 5.67) |
| External validation ICA | AP | 182 / 64 | 13.65 (10.36 to 17.96) | 3.51 (2.88 to 4.33) |
| External validation ICA | Lateral | 171 / 62 | 35.80 (30.18 to 42.48) | 7.11 (6.17 to 8.09) |

Scores are patient-level means with runs averaged within each patient before equal weighting of patients. Parentheses give 95% percentile confidence intervals from 2,000 patient-bootstrap samples. Boundary distances are measured on the 512 × 512 annotation grid. Figs. S2 and S3 show the individual observations and population summaries.

**Supplementary Table S9. ResNet50 segmentation sensitivity and precision**

| Cohort and task | View | Runs / patients | Sensitivity (95% CI) | Precision (95% CI) |
|---|---|---|---|---|
| Internal validation Hemisphere | AP | 232 / 59 | 0.965 (0.960 to 0.970) | 0.968 (0.963 to 0.972) |
| Internal validation Hemisphere | Lateral | 256 / 60 | 0.973 (0.969 to 0.977) | 0.970 (0.967 to 0.974) |
| Internal validation ICA | AP | 232 / 59 | 0.964 (0.954 to 0.974) | 0.479 (0.461 to 0.496) |
| Internal validation ICA | Lateral | 256 / 60 | 0.941 (0.925 to 0.955) | 0.459 (0.438 to 0.480) |
| External validation Hemisphere | AP | 182 / 64 | 0.960 (0.949 to 0.968) | 0.961 (0.957 to 0.966) |
| External validation Hemisphere | Lateral | 171 / 62 | 0.959 (0.951 to 0.965) | 0.972 (0.965 to 0.978) |
| External validation ICA | AP | 182 / 64 | 0.951 (0.935 to 0.965) | 0.485 (0.467 to 0.501) |
| External validation ICA | Lateral | 171 / 62 | 0.945 (0.919 to 0.966) | 0.404 (0.383 to 0.423) |

Scores are patient-level means with runs averaged within each patient before equal weighting of patients. Parentheses give 95% percentile confidence intervals from 2,000 patient-bootstrap samples. Figs. S2 and S3 show the individual observations and population summaries.

### Supplementary Table S10. Automatic versus manual agreement by cohort and projection

| View and metric | Runs / patients | Automatic mean (SD) | Manual mean (SD) | Bias (95% CI) | 95% LoA | MAE (95% CI) |
|---|---|---|---|---|---|---|
| **Internal validation** | | | | | | |
| AP MTT | 114 / 47 | 3.43 (0.93) | 3.52 (0.91) | −0.09 (−0.18 to 0.00) | −1.03 to 0.86 | 0.35 (0.30 to 0.41) |
| AP Tmax | 115 / 47 | 3.04 (0.96) | 2.98 (0.93) | 0.06 (0.02 to 0.11) | −0.41 to 0.53 | 0.16 (0.12 to 0.20) |
| Lateral MTT | 109 / 44 | 3.35 (0.90) | 3.18 (0.87) | 0.17 (0.10 to 0.24) | −0.49 to 0.83 | 0.30 (0.25 to 0.34) |
| Lateral Tmax | 109 / 44 | 2.87 (0.81) | 2.68 (0.75) | 0.19 (0.16 to 0.24) | −0.14 to 0.53 | 0.20 (0.17 to 0.25) |
| **External validation** | | | | | | |
| AP MTT | 84 / 40 | 3.59 (0.96) | 3.79 (0.88) | −0.19 (−0.31 to −0.08) | −1.16 to 0.77 | 0.39 (0.31 to 0.47) |
| AP Tmax | 84 / 40 | 2.85 (0.82) | 2.80 (0.74) | 0.06 (−0.05 to 0.18) | −0.91 to 1.02 | 0.23 (0.15 to 0.35) |
| Lateral MTT | 81 / 44 | 3.49 (0.82) | 3.34 (0.78) | 0.15 (0.05 to 0.25) | −0.69 to 0.98 | 0.32 (0.25 to 0.40) |
| Lateral Tmax | 81 / 44 | 2.77 (1.03) | 2.52 (0.59) | 0.24 (0.10 to 0.46) | −1.43 to 1.92 | 0.31 (0.17 to 0.51) |

Bias is automatic minus manual, using each pipeline’s own surviving mask and finite, strictly positive metric values. Runs are weighted equally; means are accompanied by sample SDs across runs. The 95% confidence intervals (CI) use 2,000 whole-patient bootstrap samples within cohort and projection. Marginal limits of agreement (LoA) are the mean paired difference ±1.96 times the sample SD of paired differences and describe individual-run dispersion rather than uncertainty in mean bias. MAE is the mean absolute paired difference. Internal and external results are separated because only external perfusion observations were withheld from model development. AP, anteroposterior; MTT, mean transit time; Tmax, time to maximum residue. All perfusion values are in seconds.

**Supplementary Table S11. Descriptive biplane agreement in both cohorts**

| Cohort and metric | Pairs / patients | Raw bias, s (95% CI) | Marginal 95% LoA, s | MAE, s (95% CI) |
|---|---|---|---|---|
| Internal validation MTT | 280 / 62 | 0.12 (0.02 to 0.22) | -1.03 to 1.28 | 0.46 (0.42 to 0.51) |
| Internal validation Tmax | 280 / 62 | 0.32 (0.19 to 0.47) | -1.96 to 2.61 | 0.46 (0.34 to 0.61) |
| External validation MTT | 220 / 61 | -0.07 (-0.18 to 0.04) | -1.37 to 1.24 | 0.48 (0.42 to 0.53) |
| External validation Tmax | 220 / 61 | 0.16 (-0.02 to 0.39) | -2.60 to 2.93 | 0.53 (0.35 to 0.75) |

These secondary descriptive summaries weight pairs equally and use the raw AP-minus-lateral sample mean and sample SD. Bias intervals use 2,000 whole-patient-bootstrap resamples.

**Supplementary Table S12. Repeated-measures biplane analyses and post hoc sensitivity**

| Cohort and subset | Metric | Pairs / patients | Bias, s (95% CI) | 95% LoA, s |
|---|---|---|---|---|
| External validation / all | MTT | 220 / 61 | −0.07 (-0.19 to 0.05) | −1.38 to 1.24 |
| External validation / all | Tmax | 220 / 61 | 0.16 (-0.08 to 0.40) | −2.65 to 2.97 |
| External validation / QC sensitivity | MTT | 216 / 60 | −0.03 (-0.13 to 0.07) | −1.18 to 1.13 |
| External validation / QC sensitivity | Tmax | 216 / 60 | 0.01 (-0.11 to 0.12) | −1.56 to 1.57 |
| Internal validation / QC sensitivity | MTT | 276 / 62 | 0.15 (0.06 to 0.24) | −0.87 to 1.18 |
| Internal validation / QC sensitivity | Tmax | 276 / 62 | 0.22 (0.11 to 0.33) | −1.02 to 1.46 |

Differences are AP minus lateral. REML models include patient and patient-day-vessel random intercepts. Limits of agreement (LoA) are bias ±1.96 times the square root of summed variance components; fixed-effect inference uses Kenward–Roger degrees of freedom. The post hoc sensitivity removes AP MTT below 1 second or AP Tmax at least 10 seconds, excluding four pairs in each cohort. MTT, mean transit time; Tmax, time to maximum residue. All values are seconds.

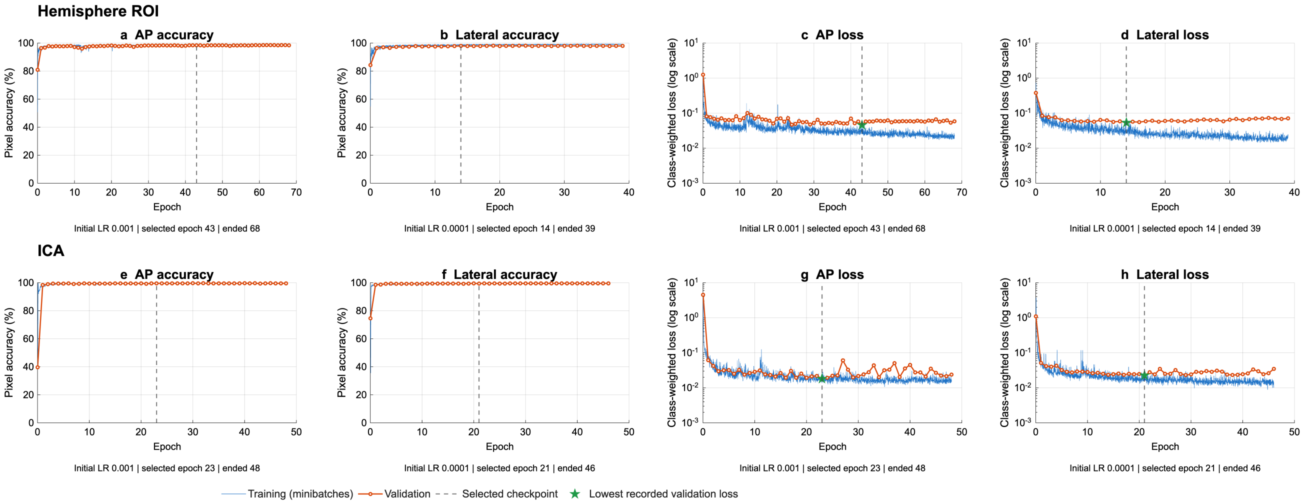


### Fig. S1 Training and validation histories of the selected ResNet50 models

Hemisphere models occupy the upper row (a–d) and ICA models the lower row (e–h). AP accuracy is shown in a/e, lateral accuracy in b/f, AP loss in c/g, and lateral loss in d/h. Loss axes are logarithmic. Blue traces show training minibatches; orange points show validation checks. Dashed lines mark selected checkpoints; green stars mark minimum validation loss. Selected epochs were 43/14 for AP/lateral hemisphere models and 23/21 for ICA models.

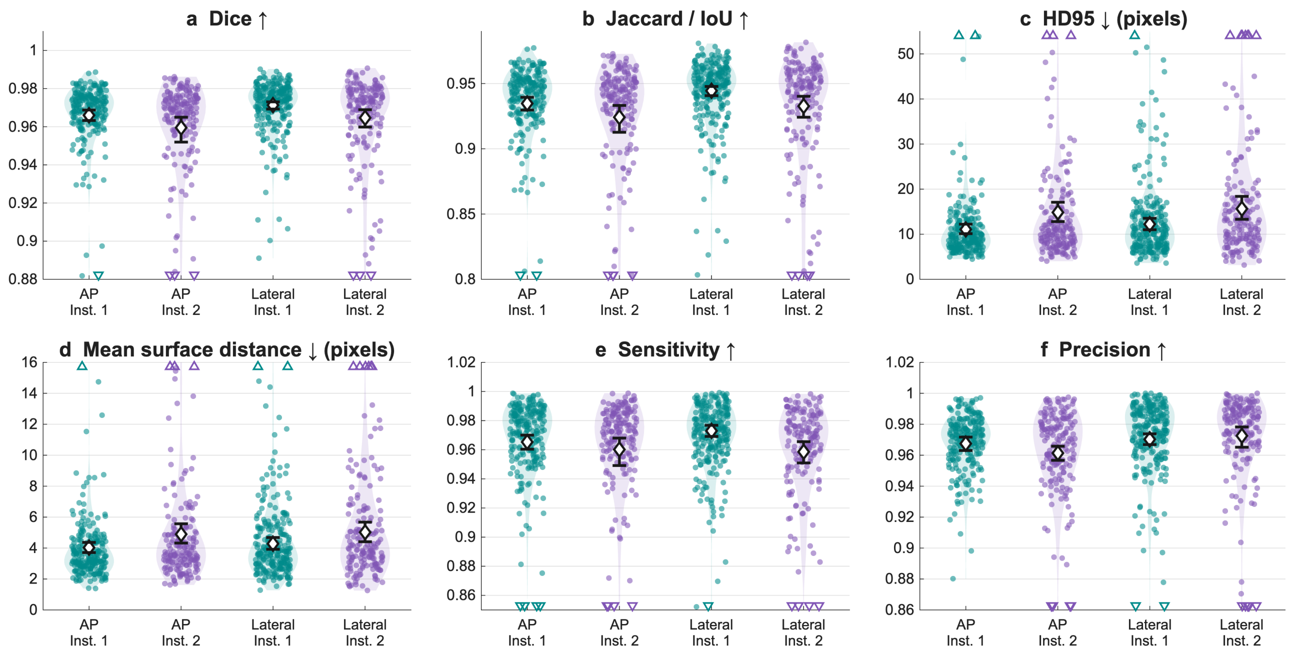


**Fig. S2 Hemisphere segmentation in both validation cohorts**

Dice (a), Jaccard (b), HD95 (c), mean symmetric boundary distance (d), sensitivity (e), and precision (f). Dots show runs; diamonds and whiskers show patient-mean scores and 95% patient-bootstrap intervals. Teal denotes internal validation (Inst. 1) and purple external validation (Inst. 2). Internal counts are 232 AP runs/59 patients and 256 lateral runs/60 patients; external counts are 182/64 and 171/62. Internal predictions are patient-held-out ResNet50 results. Triangles mark off-scale observations. Up and down arrows denote the metric direction indicating improvement in model performance.

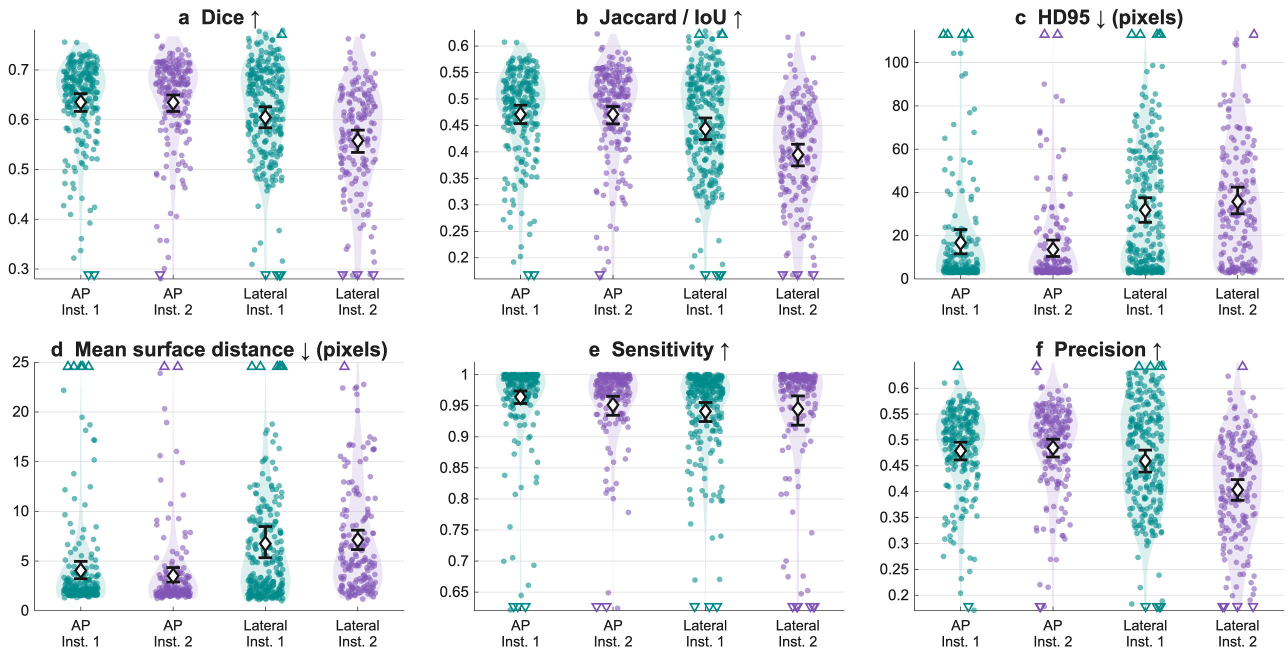


**Fig. S3 ICA segmentation in both validation cohorts**

Panels, cohort colors, sample sizes, and summaries follow Fig. S2 with Dice (a), Jaccard (b), HD95 (c), mean symmetric boundary distance (d), sensitivity (e), and precision (f). Dots show runs; diamonds and whiskers show patient-mean scores and 95% patient-bootstrap intervals. ICA masks are compared with radius-two dilated manual centerlines, using automatic hemisphere guidance in both cohorts. Up and down arrows denote the metric direction indicating improvement in model performance.

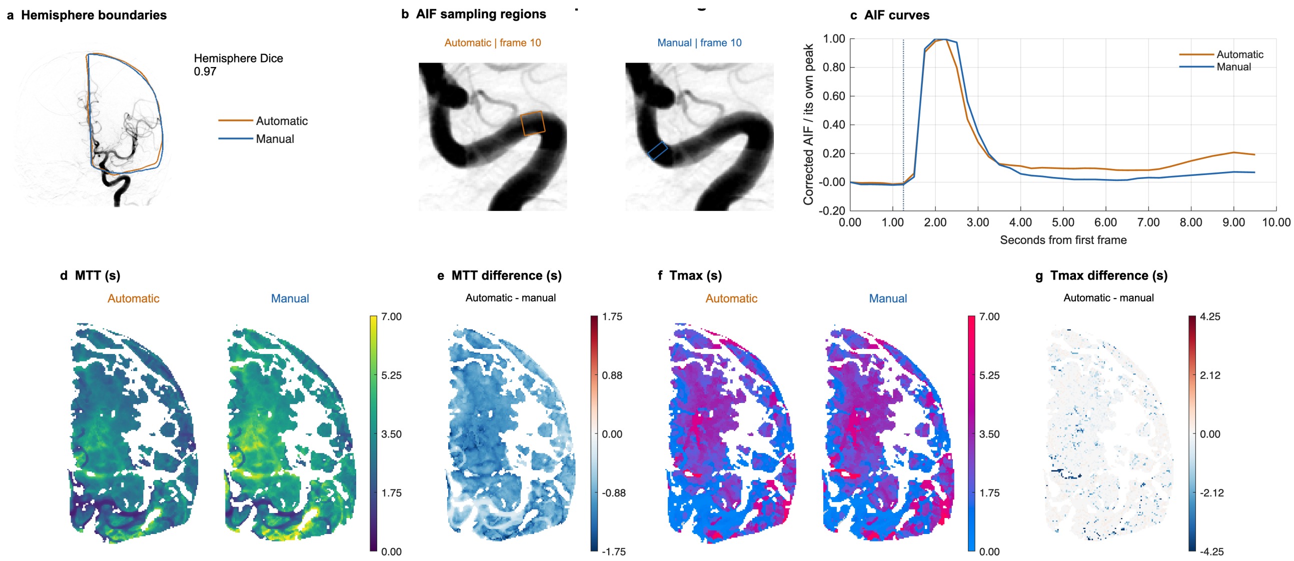


### Fig. S4 Poor automatic–manual agreement in an external AP example

This example shows a relatively poor automatic-pipeline result in comparison to manual-processing. Cases were ranked within cohort and projection by the larger absolute automatic–manual acquisition-mean difference in MTT or Tmax. The selected case lies near the 90th percentile so that approximately 90% of cases had smaller disagreement. Panels follow main-text Fig. 6: hemisphere boundaries (a), solid AIF rectangles (b), normalized AIF curves (c), paired MTT/Tmax maps (d/f), and automatic-minus-manual differences (e/g). Orange denotes automatic and blue manual processing.

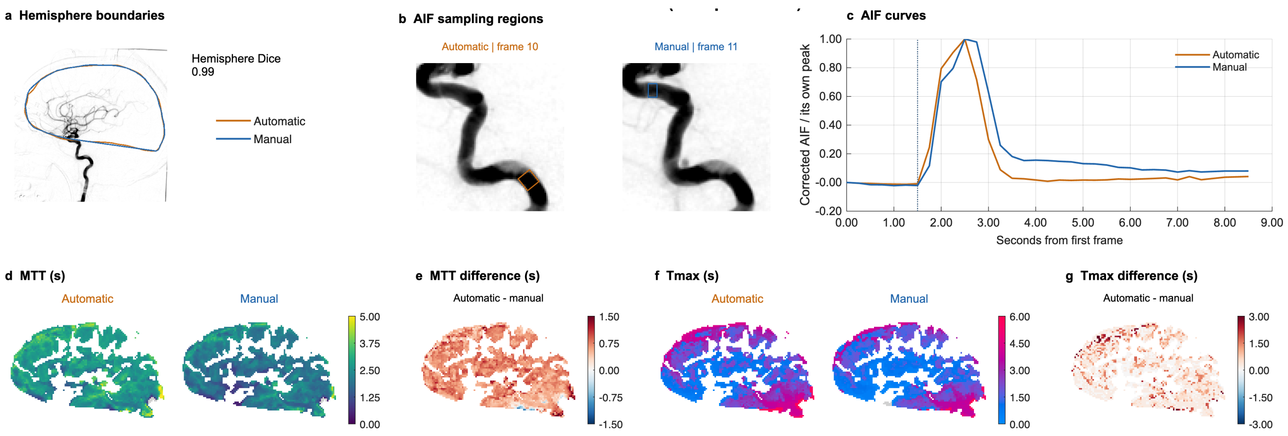


### Fig. S5 Poor automatic–manual agreement in an external lateral example

This example shows poor agreement between automatic-pipeline processing and manual processing in the lateral view. As in Fig. S4, cases were ranked within cohort and projection by the larger absolute automatic–manual acquisition-mean difference in MTT or Tmax. As in Fig. S4, hemisphere boundaries (a), solid AIF rectangles (b), normalized AIF curves (c), paired MTT/Tmax maps (d/f), and automatic-minus-manual differences (e/g). Orange denotes automatic and blue manual processing.

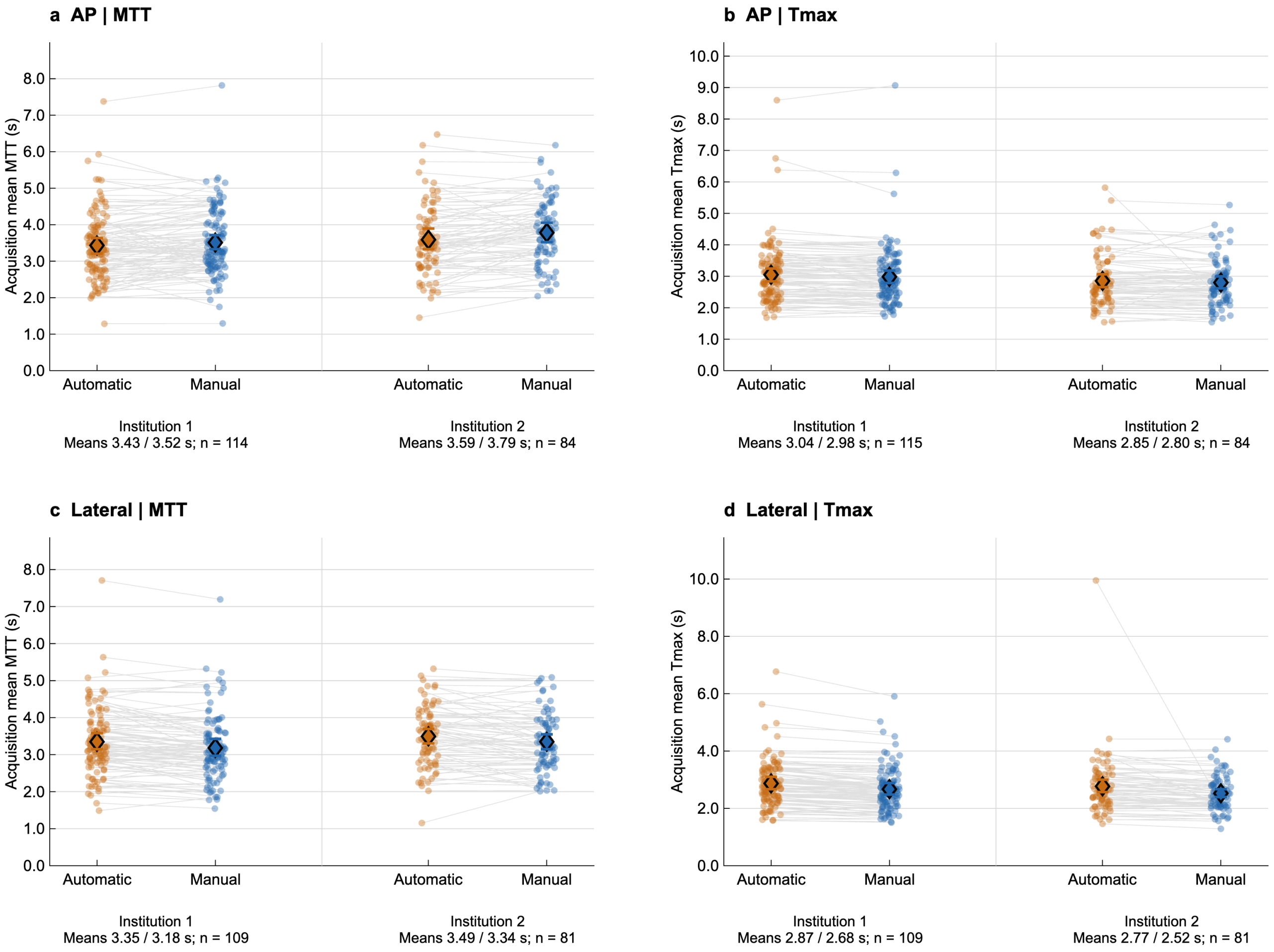


**Fig. S6 Automatic versus manual-input perfusion in both cohorts**

AP MTT (a), AP Tmax (b), lateral MTT (c), and lateral Tmax (d). Orange and blue dots show automatic and manual acquisition means, joined within runs. Diamonds and whiskers show means and 95% whole-patient-bootstrap intervals. Inst. 1 denotes internal validation and Inst. 2 external validation. Internal AP MTT includes 114 runs, AP Tmax 115, and lateral metrics 109; external AP and lateral metrics include 84 and 81 runs. Internal perfusion analyses include development patients but is shown for context.
